# Folic acid-containing supplement use among females aged 15-55 in the Canadian Community Health Survey 2015-2018

**DOI:** 10.1101/2025.01.21.25320917

**Authors:** Kathryn E. Hopperton, Lidia Loukine, The Minh Luong, Loan Nguyen, Jesse Bertinato, Yvette Bonvalot, Marcia Cooper, Wei Luo, Amanda J. MacFarlane, Jennifer McCrea, Carley Nicholson, Huma Rana, Kelsey Vercammen, Jane Yuan, Shawn Brule, Hope A. Weiler

**Author notes:** **Corresponding author:** Kathryn Hopperton, 251 Sir Frederick Banting Driveway, Ottawa, Ontario K1A 0K9 1. **Author’s last names for indexing:** Hopperton, Loukine, Luong, Nguyen, Bertinato, Bonvalot, Cooper, Luo, MacFarlane, McCrea, Nicholson, Rana, Vercammen, Yuan, Brule, Weiler. **Disclaimers:** none.

## Abstract

**Introduction:** In Canada, those who are or who could become pregnant are recommended to consume a daily multivitamin containing 400 µg of folic acid to help prevent neural tube defects.

**Objectives:** To report the prevalence and determinants of folic acid-containing supplement use among females of childbearing age in Canada.

**Methods:** Data were combined from cycles 2015/16 and 2017/18 of the maternal experiences module of the cross-sectional Canadian Community Health Survey, which was completed by females aged 15-55 years. Representative weighted estimates (means/percentages, 95% CI) were generated for folic acid-containing supplement use among all pregnant, non-pregnant, and lactating respondents. For those who had given birth in the preceding 5 years, estimates were also generated for supplement use in the 3 months before and first 3 months of their most recent pregnancy, and pre-pregnancy awareness of the link between folic acid and some birth defects. We examined associations with sociodemographic factors using multivariable logistic regression.

**Results:** Overall, 16.5% (15.9-17.0%) of non-pregnant, 80.3% (77.1-83.5%) of pregnant, and 58.4% (54.8-61.9%) of lactating females aged 15-55 reported using a folic acid-containing supplement. Among those who had given birth in the preceding 5 years, 63.7% (62.2-65.1%) consumed a folic acid-containing supplement in the 3 months prior to pregnancy, while 89.9% (88.8-90.9%) did so during the first trimester. Lower prevalence of supplement use before or during pregnancy was reported among the 23.7% (22.4-25.1%) of respondents unaware of the relationship between folic acid and birth defects. Younger age, single marital status, lower educational attainment, income below the median, and smoking were associated with lower odds of awareness or supplement use.

**Conclusion:** While most females living in Canada reported using folic-acid containing supplements prior to and during pregnancy, use of these supplements among non-pregnant females of childbearing age is low, and sociodemographic inequalities exist.

## INTRODUCTION

Inadequate folate status in the periconceptional period is associated with a higher risk of neural tube defects (NTDs) among offspring (1–3). NTDs are a group of serious congenital conditions arising from incomplete closure of the neural tube between weeks 3 and 4 of gestation (4). Data from Canada and the United States suggests that 40-50% of pregnancies are unplanned, with a higher proportion of unplanned pregnancies among those in younger age groups (5, 6). The folate status of those who could become pregnant is therefore a significant public health concern for the prevention of NTDs, whether a pregnancy is planned or not.

To meet the needs of a diverse population, a multipronged approach to achieving adequate folate status as primary prevention of NTDs is likely necessary. Randomized controlled trials in the 1990s demonstrated substantial reductions in the risk of NTD occurrence and recurrence among participants randomized to consume a supplement containing folic acid in the period around conception (1, 2). Based largely on these findings, many countries, including Canada and the United States, implemented mandatory fortification of the food supply with folic acid (3, 7). Following the introduction of folic acid fortification in Canada in 1998, the prevalence of NTDs declined from 1.58 to 0.86 per 1000 births, a 46% reduction (95% CI: 40-51) (8). Other countries have seen similar declines in NTD prevalence following the introduction of fortification, with a 2019 meta-analysis of 8 studies and over 19 million births or still births reporting a 41% decrease (OR 0.59 [95% CI: 0.49-0.70]) (9).

Despite this success, folic acid fortification alone is not sufficient to minimize population risk of NTDs. Nationally representative data from the 2015 Canadian Community Health Survey (CCHS) - Nutrition estimated that over 20% females of reproductive age have usual folate intakes below the estimated average requirement (EAR) from food sources alone (10, 11). Complementing this, representative national biomonitoring data from cycle 1 of the Canadian Health Measures Survey (2007–2009) showed that 22% of females aged 15-45 had red blood cell folate concentrations <906 nmol/L, which is the level above which maximal NTD risk reduction is expected (12). Subsequent analyses have suggested that this may be an underestimation (13). Post-fortification data from the National Health and Nutrition Examination Survey (NHANES) suggests that a similar proportion of U.S women have suboptimal status (22.8% [95% CI: 21.1-24.6]) (14). Canadian public health agencies therefore recommend that anyone who could become pregnant consume a daily multivitamin containing 400 µg of folic acid, in addition to consuming a healthy diet that includes other sources of folate, to reduce their risk of a pregnancy affected by a NTD (4, 15).

Nationally representative data on folic acid-containing supplement use by females of childbearing age is collected annually in the CCHS, but the last published estimates are from 2006-7 post-censal Maternity Experiences Survey (16–18). The primary objective of our study was to provide updated estimates of the prevalence of folic acid-containing supplement use among pregnant and non-pregnant females aged 15-55 living in Canada, and of awareness of the link between folic acid and NTDs. Our secondary objective was to explore sociodemographic, geographic and health characteristics associated with supplement use that could help to tailor future public health initiatives within Canada and other countries with similar recommendations. We used two recent cycles of the CCHS (covering 2015-18) and examined folic acid-containing supplement use both among those who reported giving birth in the preceding 5 years, and among all females of childbearing age separated into groups of public health interest (non-pregnant, pregnant, and lactating).

## SUBJECTS AND METHODS

### The Canadian Community Health Survey Cycles 2015-2018

The CCHS is a cross-sectional survey conducted annually by Statistics Canada, a Canadian federal government organization, in partnership with Health Canada and the Public Health Agency of Canada.

The survey was voluntary, and data collection was conducted according to the Statistics Act of Canada. It provides representative regional estimates of the health status, health determinants and healthcare utilization of people living in Canada every two years. Participants include those aged ≥ 12 years living in any of Canada’s provinces or territories, with the following exclusions (representing <3% of the population): those in the provinces living on reserves or in other Aboriginal settlements, full-time members of the Canadian forces, those living in institutions, children aged 12-17 years in foster care, and certain regions of northern Quebec. We selected cycles 2015/16 and 2017/18 for analysis as these were the most recent cycles with a consistent sampling frame (changed after 2019) that were not impacted by changes in sampling methodology and response rate that occurred due to the COVID-19 pandemic. The response rate for cycle 2015/16 was 59.5% overall, while for 2017/18 it was 58.8%, with some geographic variation (19, 20). Surveys were conducted by trained interviewers in-person or over the telephone (for the maternal experiences module 27% and 73%, respectively).

### Folic acid-containing supplement use

Data on folic acid-containing supplement use was obtained from the Maternal Experiences module, which was administered as core content to respondents who reported being female and between 15-55 years of age at the time of the survey. Details of the specific questions and their administration are available in the **Supplemental Materials**. Respondents answered questions about their supplement use at the time of the survey (referred to henceforth as “current supplement use”), which we examined separately within participants who reported being currently pregnant, currently lactating but not pregnant (henceforth called “lactating”), and not currently pregnant. Respondents who reported having given birth in the last five years answered additional questions about their use of folic acid-containing supplements in the three months prior to their most recent pregnancy, and in the first three months of their most recent pregnancy. For ease of reporting, we refer to this period of supplementation as “peri-conceptional”. If they answered “Yes” to either question about folic acid-containing supplement use peri-conceptionally, they were asked a follow-up question regarding frequency. Respondents were also asked about their awareness of the link between folic acid and birth defects. Our final analytical sample included those who answered “yes” or “no” to these questions.

### Other variables

We examined folic acid-containing supplement use by categories of respondents within sociodemographic, geographic, and health attributes available in CCHS identified *a priori* based on previous literature and knowledge of determinants of supplement use, folate status, or healthcare access. Details of the derivation and categorization of these variables is available in the **Supplemental Materials.** The variables included: age, highest level of education completed, household size-adjusted income, source of household income, marital status, immigration status, self-reported racial group or indigenous identity, province or territory of residence, region of residence, urban or rural location of residence, food security, BMI, daily consumption of fruits and vegetables, monthly consumption of green vegetables, smoking, alcohol use, and frequency of general physical checkup. Because folic acid-containing supplement use in the peri-conception period was assessed retrospectively, we additionally examined time since most recent birth (0 to 5 years) for these outcomes to assess the impact of recall bias. For all questions, missing data from responses “don’t know”, refusal or not stated were combined.

### Statistical Analysis

All statistical analyses were carried out in SAS version 9.4 using the CCHS shared file with data from respondents who consented to data sharing with Health Canada and the Public Health Agency of Canada (93.6% for cycles 2015/16 and 93.1% for cycles 2017/18). Survey sampling weights from Statistics Canada were applied to all estimates of proportion to account for the sampling design and probability of selection so that study results can be considered representative of the population of Canada (19, 20). We used the bootstrap method to calculate 95% confidence intervals of all estimates, applying 1000 bootstrap weights produced by Statistics Canada. Data from different cycles were combined with adjusted cycle weights per established Statistics Canada methodology (21).

We fit regression models for the use of a folic-acid containing supplement or awareness of the link between folic acid and birth defects between categories of the sociodemographic, geographic and health variables in both a univariate model and in multivariable models adjusting for significant factors from the univariate analyses. Education was colinear with income, while region of residence overlapped in definition with province or territory of residence, so these sociodemographic factors were examined in separate models. The models for income variables, which were only recorded in 2017-18, were limited to respondents from this latter survey. Frequency of general physical checkup was included as optional content in the 2015/16 survey only and was answered by <50% of respondents, so this variable was examined in the univariate model only. The association between outcomes and factors was assessed using ANOVA, with p-value<0.05 to denote statistical significance. Post-hoc pairwise comparisons were conducted for factors with significant global associations, with odds ratios reported vs a pre-specified reference group. In line with Statistics Canada reporting requirements, we suppressed results for groups with a sample size <10 or with a CV >33.3% as unacceptable for reporting (replaced with the letter “F”), and flagged groups with a coefficient of variation (CV) of >16.6-33.2% to be interpreted with caution with the letter “E” (19, 20).

## RESULTS

A total of 59,345 females aged 15-55 years completed the Maternal Experiences module, including 29,635 from the 2015-16 cycle, and 29,710 from the 2017-18 cycle. The participant flow diagram is shown in **Figure 1**. Current supplement use was stratified by pregnancy and lactation status: not pregnant or lactating, (n=53,559, mean age 36.1 [95% CI: 36.0, 36.2] years), lactating, (n=2069, mean age 32.3 [95% CI 32.0, 32.6], and pregnant (n=1,676, mean age 31.7 [95% CI: 31.2, 32.2] years). Among those who reported having given birth in the last five years, we assessed folic acid-containing supplement use in the three months prior to their most recent pregnancy (n=10,326, mean age at delivery 30.8 [95% CI: 30.6, 30.9] years), and during the first 3 months of their most recent pregnancy (n=10,329, mean age at delivery 30.8 [95% CI: 30.6, 30.9] years). We also examined awareness of the link between folic acid and some birth defects prior to pregnancy in this group (n=10,384, mean age at delivery 30.7 [95% CI: 30.6, 30.9] years).

**Figure 1.**
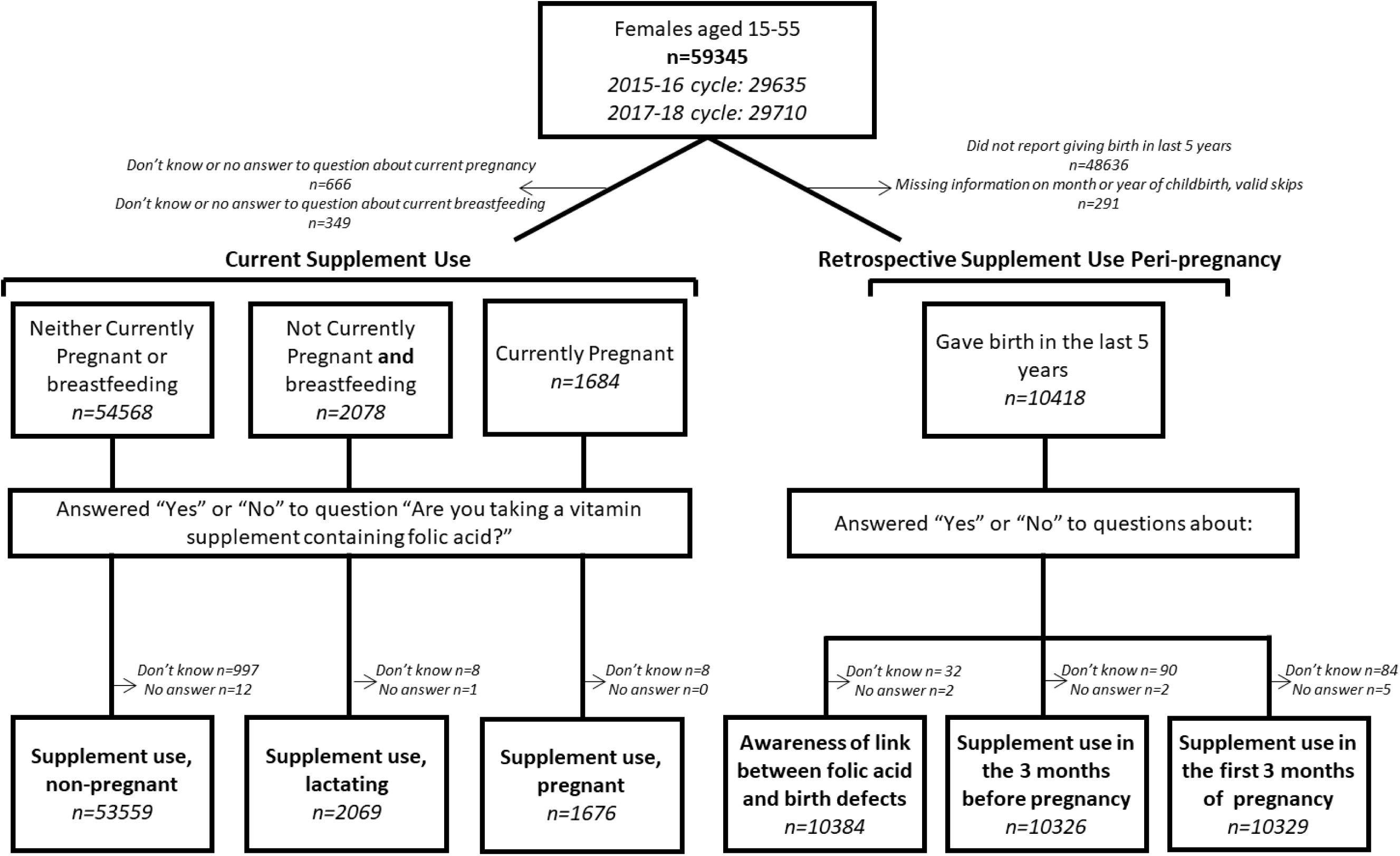
Participant Flow.

### Prevalence of folic acid-containing supplement use

Overall, 16.5% (95% CI: 15.9, 17.0) of females of childbearing age living in Canada who were not pregnant or lactating reported currently taking a vitamin supplement containing folic acid (**Table 1)**. The proportion was highest in Alberta (21.7% [95% CI: 20.2, 23.2) and lowest in Nunavut 8.1% [95% CI: 3.5, 12.8] E) and Quebec (7.3% (95% CI: 6.5, 8.0). The proportion was higher among non-pregnant respondents who were lactating (58.4%, [95% CI 54.8, 61.9]) (**Table 2**) and among those who reported being currently pregnant, (80.3% [95% CI: 77.1, 83.5]) (**Table 3**). Among respondents who had given birth in the preceding 5 years, 76.3% (95% CI: 74.9, 77.6) reported that they had been aware prior to their pregnancy that taking folic acid can help prevent some birth defects **(Table 4)**. Awareness was highest in Prince Edward Island (84.1%, [95% CI: 77.6, 90.7]) and lowest in Nunavut (18.1% [95% CI: 10.6, 25.5 E]. In the 3 months prior to becoming pregnant, 63.7% (95% CI: 62.2-65.1) of respondents reported that they had taken a folic acid-containing supplement (**Table 5)**, with the highest proportions among respondents from Quebec (68.5% [95% CI: 65.7, 71.2]) and Prince Edward Island (68.1% [95% CI: 59.0, 77.3]), and the lowest proportion in Newfoundland and Labrador (49.1% [95% CI: 40.4, 57.8]) and Nunavut (22.4% [95% CI: 13.8, 30.9 E]). During the first 3 months of their most recent pregnancy, 89.9% reported having taken a folic acid-containing supplement (95% CI: 88.8, 90.9), with the highest proportions in Prince Edward Island (97.1% [95% CI: 94.3, 99.9]) and British Columbia (94.3% [95% CI: 92.4, 96.2]), and the lowest in Manitoba (84.0% [95% CI: 79.7, 88.2]) and Nunavut (54.9% [95% CI: 43.7, 66.1]) (**Table 6)**.

**Table 1.** Folic acid-containing supplement use by among females aged 15-55 who are neither pregnant or lactating^1^.

| Subgroup | Sample Size<sup>2</sup> | Supplement Users (% [95% CI]) | Supplement Non-users (% [95% CI]) | OR Unadjusted (95% CI) | OR Adjusted (95% CI) |
| --- | --- | --- | --- | --- | --- |
| Are you taking a vitamin supplement containing folic acid?" among those who report neither being currently pregnant or breastfeeding | 53559 | 16.5 (15.9, 17.0) | 83.5 (83.0, 84.1) | n/a | n/a |
| **Current age** |  |  |  |  |  |
| <=30 | 15938 | 11.0 (10.2, 11.8) | 89.0 (88.2, 89.8) | **0.52 (0.48, 0.57)** | **0.70 (0.63, 0.79)** |
| 31-55 | 37621 | 19.2 (18.5, 19.9) | 80.8 (80.1, 81.5) | Ref | Ref |
| **Highest level of education completed<sup>3</sup>** |  |  |  |  |  |
| ≤ High school diploma | 18699 | 11.0 (10.2, 11.7) | 89.0 (88.3, 89.8) | **0.59 (0.54, 0.66)** | **0.69 (0.62, 0.77)** |
| Postsecondary diploma | 19516 | 17.2 (16.3, 18.1) | 82.8 (81.9, 83.7) | Ref | Ref |
| Bachelor's or higher | 14896 | 21.5 (20.4, 22.6) | 78.5 (77.4, 79.6) | **1.32 (1.20, 1.44)** | **1.22 (1.11, 1.34)** |
| **Household size-adjusted household income<sup>4</sup>** |  |  |  |  |  |
| < median (42146.57) | 11071 | 14.7 (13.6, 15.8) | 85.3 (84.2, 86.4) | n.s | n/a |
| >= median (42146.57) | 15755 | 16.1 (15.2, 17.0) | 83.9 (83.0, 84.8) |  |  |
| **Marital status** |  |  |  |  |  |
| Married, Common-law | 27576 | 20.6 (19.8, 21.4) | 79.4 (78.6, 80.2) | Ref | Ref |
| Other (single, widowed, separated, divorced) | 25904 | 11.4 (10.8, 12.1) | 88.6 (87.9, 89.2) | **0.50 (0.46, 0.54)** | **0.61 (0.56, 0.67)** |
| **Immigration status** |  |  |  |  |  |
| Immigrated ≤5 years | 1772 | 20.3 (17.1, 23.5) | 79.7 (76.5, 82.9) | **1.38 (1.12, 1.69)** | 0.96 (0.76, 1.21) |
| Immigrated >5 years | 7244 | 18.6 (17.1, 20.0) | 81.4 (80.0, 82.9) | **1.23 (1.11, 1.37)** | 0.90 (0.79, 1.03) |
| *Non-immigrant (born in Canada)* | 44194 | 15.6 (15.1, 16.1) | 84.4 (83.9, 84.9) | Ref | Ref |
| **Self-reported race or Indigenous identity<sup>5</sup>** |  |  |  |  |  |
| Black | 1043 | 19.4 (15.9, 23.0) | 80.6 (77.0, 84.1) | **1.29 (1.02, 1.63)** | **1.65 (1.28, 2.12)** |
| East/Southeast Asian | 2996 | 15.8 (13.7, 17.9) | 84.2 (82.1, 86.3) | 1.00 (0.85, 1.18) | 0.89 (0.73, 1.07) |
| First Nations | 1950 | 13.8 (11.5, 16.2) | 86.2 (83.8, 88.5) | 0.86 (0.70, 1.05) | 0.95 (0.77, 1.17) |
| Inuit | 492 | 9.3 (4.8, 13.7) E | 90.7 (86.3, 95.2) | **0.55 (0.31, 0.97)** | 0.71 (0.37, 1.36) |
| Latin American | 585 | 13.2 (9.7, 16.8) | 86.8 (83.2, 90.3) | 0.82 (0.60, 1.12) | 0.85 (0.61, 1.19) |
| Métis | 1540 | 15.7 (12.9, 18.4) | 84.3 (81.6, 87.1) | 0.99 (0.80, 1.23) | 1.01 (0.80, 1.27) |
| Middle Eastern | 661 | 18.8 (14.2, 23.3) | 81.2 (76.7, 85.8) | 1.24 (0.92, 1.67) | 1.32 (0.95, 1.83) |
| Multiple Indigenous | 134 | 10.2 (4.0, 16.4) E | 89.8 (83.6, 96.0) | 0.61 (0.30, 1.23) | 0.79 (0.39, 1.60) |
| Not stated | 439 | 16.1 (10.6, 21.7) E | 83.9 (78.3, 89.4) | 1.03 (0.68, 1.56) | 0.81 (0.46, 1.45) |
| Other or multiple races | 1524 | 19.0 (15.8, 22.1) | 81.0 (77.9, 84.2) | **1.25 (1.02, 1.55)** | **1.39 (1.11, 1.73)** |
| South Asian | 1361 | 25.1 (21.4, 28.8) | 74.9 (71.2, 78.6) | **1.79 (1.47, 2.19)** | **1.66 (1.32, 2.08)** |
| *White* | 40834 | 15.7 (15.2, 16.3) | 84.3 (83.7, 84.8) | Ref | Ref |
| **Province or territory of residence** |  |  |  |  |  |
| Alberta | 7115 | 21.7 (20.2, 23.2) | 78.3 (76.8, 79.8) | **1.21 (1.08, 1.35)** | **1.39 (1.20, 1.59)** |
| British Columbia | 6898 | 21.5 (20.0, 23.0) | 78.5 (77.0, 80.0) | **1.20 (1.07, 1.34)** | **1.40 (1.22, 1.61)** |
| Manitoba | 2689 | 18.5 (16.4, 20.6) | 81.5 (79.4, 83.6) | 0.99 (0.84, 1.16) | **1.21 (>1.00, 1.46)** |
| New Brunswick | 1557 | 12.1 (10.1, 14.2) | 87.9 (85.8, 89.9) | **0.60 (0.50, 0.73)** | **0.71 (0.57, 0.88)** |
| Newfoundland and Labrador | 1525 | 11.4 (9.4, 13.5) | 88.6 (86.5, 90.6) | **0.56 (0.45, 0.70)** | **0.61 (0.48, 0.76)** |
| Northwest Territories | 561 | 17.9 (13.8, 22.1) | 82.1 (77.9, 86.2) | 0.95 (0.70, 1.29) | 1.25 (0.88, 1.77) |
| Nova Scotia | 2250 | 14.6 (12.4, 16.7) | 85.4 (83.3, 87.6) | **0.74 (0.62, 0.89)** | 0.88 (0.72, 1.07) |
| Nunavut | 481 | 8.1 (3.5, 12.8) E | 91.9 (87.2, 96.5) | **0.39 (0.21, 0.73)** | 0.67 (0.27, 1.64) |
| *Ontario* | 15600 | 18.7 (17.6, 19.7) | 81.3 (80.3, 82.4) | Ref | Ref |
| Prince Edward Island | 882 | 12.0 (9.5, 14.5) | 88.0 (85.5, 90.5) | **0.60 (0.47, 0.76)** | **0.71 (0.55, 0.92)** |
| Quebec | 11170 | 7.3 (6.5, 8.0) | 92.7 (92.0, 93.5) | **0.34 (0.30, 0.39)** | **0.37 (0.32, 0.43)** |
| Saskatchewan | 2310 | 20.3 (18.0, 22.7) | 79.7 (77.3, 82.0) | 1.11 (0.95, 1.30) | **1.36 (1.14, 1.63)** |
| Yukon | 521 | 17.7 (13.8, 21.7) | 82.3 (78.3, 86.2) | 0.94 (0.71, 1.24) | 0.99 (0.74, 1.31) |

1 All characteristics refer to current status at the time the survey was conducted, except where noted otherwise. Multivariable models included all factors significantly associated with folic acid-containing supplement use in the univariate model; factors whose global association was not significant were excluded and denoted with n.s. Separate models were run for income and education, and for province/territory of residence and region of residence to avoid collinearity. Bold indicates significant differences ( $p < 0.05$ ) from the reference group after adjustment for multiple comparisons. Superscript E indicates that values should be interpreted with caution due to a wide CV (16.6-33.3%), while estimates replaced by an F are suppressed due to low sample size ( $< 10$ ) and/or a CV $\geq 33.3\%$ .
2 Sample sizes refer to those who answered either yes or no. Sample sizes and proportions may not add to 100%/the full sample size because some responses were do not know or refuse to answer, or because question was not included for all cycles/and/or provinces/territories as specified below.
3 Categorized as < high school, postsecondary [trade, college, CEGEP or other non-university or university certificate or diploma below the bachelor's level], or university [bachelor's degree or certificate, diploma or degree above the bachelor's level].
4 Total household income before taxes adjusted for household size (income/square root of size)(37). Income available in this format only for 2017/18 cycle.
5 Self-reported race or Indigenous identity categorized according to Canadian Institute for Health Information Draft Guidance on the Use of Standards for Race-Based and Indigenous Identity Data Collection and Health Reporting in Canada(38).

**Table 2.** Folic acid-containing supplement use by lactating females aged 15-55^1^.

| Subgroup | Sample Size<sup>2</sup> | Supplement Users (% [95% CI]) | Supplement Non-users (% [95% CI]) | OR Unadjusted (95% CI) | OR Adjusted (95% CI) |
| --- | --- | --- | --- | --- | --- |
| Are you taking a vitamin supplement containing folic acid?" among those who report not being pregnant and currently breastfeeding | 2069 | 58.4 (54.8, 61.9) | 41.6 (38.1, 45.2) | n/a | n/a |
| **Current age** |  |  |  |  |  |
| <=30 | 641 | 57.3 (50.8, 63.8) | 42.7 (36.2, 49.2) | n.s | n/a |
| 31-55 | 1428 | 58.8 (54.8, 62.8) | 41.2 (37.2, 45.2) |  |  |
| **Highest level of education completed<sup>3</sup>** |  |  |  |  |  |
| ≤ High school diploma | 446 | 52.2 (44.6, 59.9) | 47.8 (40.1, 55.4) | n.s | n/a |
| Postsecondary diploma | 678 | 58.2 (52.1, 64.2) | 41.8 (35.8, 47.9) |  |  |
| Bachelor's or higher | 938 | 60.6 (55.5, 65.7) | 39.4 (34.3, 44.5) |  |  |
| **Household size-adjusted household income<sup>4</sup>** |  |  |  |  |  |
| < median (42146.57) | 487 | 51.6 (44.4, 58.8) | 48.4 (41.2, 55.6) | Ref | Ref |
| >= median (42146.57) | 531 | 63.1 (56.6, 69.6) | 36.9 (30.4, 43.4) | **1.61 (1.08, 2.38)** | 1.48 (<1.00, 2.20) |
| **Marital status** |  |  |  |  |  |
| Married, Common-law | 1862 | 59.8 (56.1, 63.6) | 40.2 (36.4, 43.9) | Ref | Ref |
| Other (single, widowed, separated, divorced) | 206 | 34.4 (24.7, 44.2) | 65.6 (55.8, 75.3) | **0.35 (0.22, 0.56)** | **0.41 (0.24, 0.70)** |
| **Immigration status** |  |  |  |  |  |
| Immigrated ≤5 years | 189 | 50.2 (38.9, 61.5) | 49.8 (38.5, 61.1) | n.s | n/a |
| Immigrated >5 years | 341 | 57.6 (49.3, 66) | 42.4 (34, 50.7) |  |  |
| *Non-immigrant (born in Canada)* | 1524 | 59.4 (55.4, 63.4) | 40.6 (36.6, 44.6) |  |  |
| **Self-reported race or Indigenous identity<sup>5</sup>** |  |  |  |  |  |
| Black | 70 | 39.6 (23.7, 55.6) E | 60.4 (44.4, 76.3) | n.s | n/a |
| East/Southeast Asian | 168 | 67.9 (58.8, 77.1) | 32.1 (22.9, 41.2) |  |  |
| First Nations | 64 | 45.3 (26.8, 63.8) E | 54.7 (36.2, 73.2) E |  |  |
| Inuit | 51 | F | 94.1 (85.2, 100.0) |  |  |
| Latin American | 29 | 53.6 (30.0, 77.2) E | 46.4 (22.8, 70.0) |  |  |
| Métis | 53 | 75.8 (59.0, 92.6) | 24.2 (7.4, 41.0) |  |  |
| Middle Eastern | 39 | 62.8 (42.9, 82.8) | 37.2 (17.2, 57.1) |  |  |
| Multiple Indigenous | 7 | 73.7 (25.1, 100.0) F | F |  |  |
| Not stated | 21 | 78.3 (53.4, 100.0) | 21.7 (0, 46.6) |  |  |
| Other or multiple races | 68 | 48.1 (29.0, 67.1) E | 51.9 (32.9, 71) |  |  |
| South Asian | 81 | 59.3 (37.9, 80.8) E | 40.7 (19.2, 62.1) |  |  |
| *White* | 1418 | 57.9 (53.9, 62.0) | 42.1 (38.0, 46.1) |  |  |
| **Province or territory of residence** |  |  |  |  |  |
| Alberta | 343 | 59.2 (52.3, 66.1) | 40.8 (33.9, 47.7) | 0.77 (0.51, 1.15) | 0.55 (0.24, 1.24) |
| British Columbia | 313 | 65.9 (58.1, 73.8) | 34.1 (26.2, 41.9) | 1.02 (0.66, 1.59) | 0.82 (0.45, 1.48) |
| Manitoba | 126 | 43.9 (31.0, 56.7) | 56.1 (43.3, 69.0) | **0.41 (0.23, 0.75)** | **0.28 (0.11, 0.70)** |
| New Brunswick | 38 | 66.0 (49.0, 82.9) | 34.0 (17.1, 51.0) | 1.02 (0.46, 2.29) | 0.69 (0.23, 2.13) |
| Newfoundland and Labrador | 24 | F | 61.2 (27.4, 94.9) E | 0.34 (0.07, 1.56) | 0.40 (0.09, 1.90) |
| Northwest Territories | 44 | 41.2 (19.0, 63.4) E | 58.8 (36.6, 81.0) E | **0.37 (0.14, 0.99)** | 0.25 (0.06, 1.03) |
| Nova Scotia | 61 | 65.3 (49.8, 80.9) | 34.7 (19.1, 50.2) E | 0.99 (0.47, 2.12) | 0.71 (0.26, 1.92) |
| Nunavut | 59 | F | 86.9 (75.3, 98.4) | **0.08 (0.02, 0.27)** | **0.08 (0.02, 0.39)** |
| *Ontario* | 548 | 65.5 (59.1, 71.9) | 34.5 (28.1, 40.9) | Ref | Ref |
| Prince Edward Island | 28 | 45.4 (23.8, 67.0) E | 54.6 (33.0, 76.2) E | 0.44 (0.16, 1.19) | **0.31 (0.10, 0.96)** |
| Quebec | 351 | 39.6 (32.3, 46.9) | 60.4 (53.1, 67.7) | **0.35 (0.23, 0.52)** | **0.25 (0.11, 0.56)** |
| Saskatchewan | 110 | 55.9 (43.3, 68.5) | 44.1 (31.5, 56.7) | 0.67 (0.37, 1.21) | 0.44 (0.18, 1.12) |
| Yukon | 24 | 60.5 (36.0, 85.0) E | 39.5 (15.0, 64.0) E | 0.81 (0.26, 2.53) | 0.73 (0.16, 3.38) |

1 All characteristics refer to current status at the time the survey was conducted, except where noted otherwise. Multivariable models included all factors significantly associated with folic acid-containing supplement use in the univariate model; factors whose global association was not significant were excluded and denoted with n.s. Separate models were run for income and education, and for province/territory of residence and region of residence to avoid collinearity. Bold indicates significant differences ( $p < 0.05$ ) from the reference group after adjustment for multiple comparisons. Superscript E indicates that values should be interpreted with caution due to a wide CV (16.6-33.3%), while estimates replaced by an F are suppressed due to low sample size ( $< 10$ ) and/or a CV $\geq 33.3\%$ .
2 Sample sizes refer to those who answered either yes or no. Sample sizes and proportions may not add to 100%/the full sample size because some responses were do not know or refuse to answer, or because question was not included for all cycles/and/or provinces/territories as specified below.
3 Categorized as < high school, postsecondary [trade, college, CEGEP or other non-university or university certificate or diploma below the bachelor's level], or university [bachelor's degree or certificate, diploma or degree above the bachelor's level].
4 Total household income before taxes adjusted for household size (income/square root of size)<sup>(37)</sup>. Income available in this format only for 2017/18 cycle.
5 Self-reported race or Indigenous identity categorized according to Canadian Institute for Health Information Draft Guidance on the Use of Standards for Race-Based and Indigenous Identity Data Collection and Health Reporting in Canada<sup>(38)</sup>.

**Table 3.**
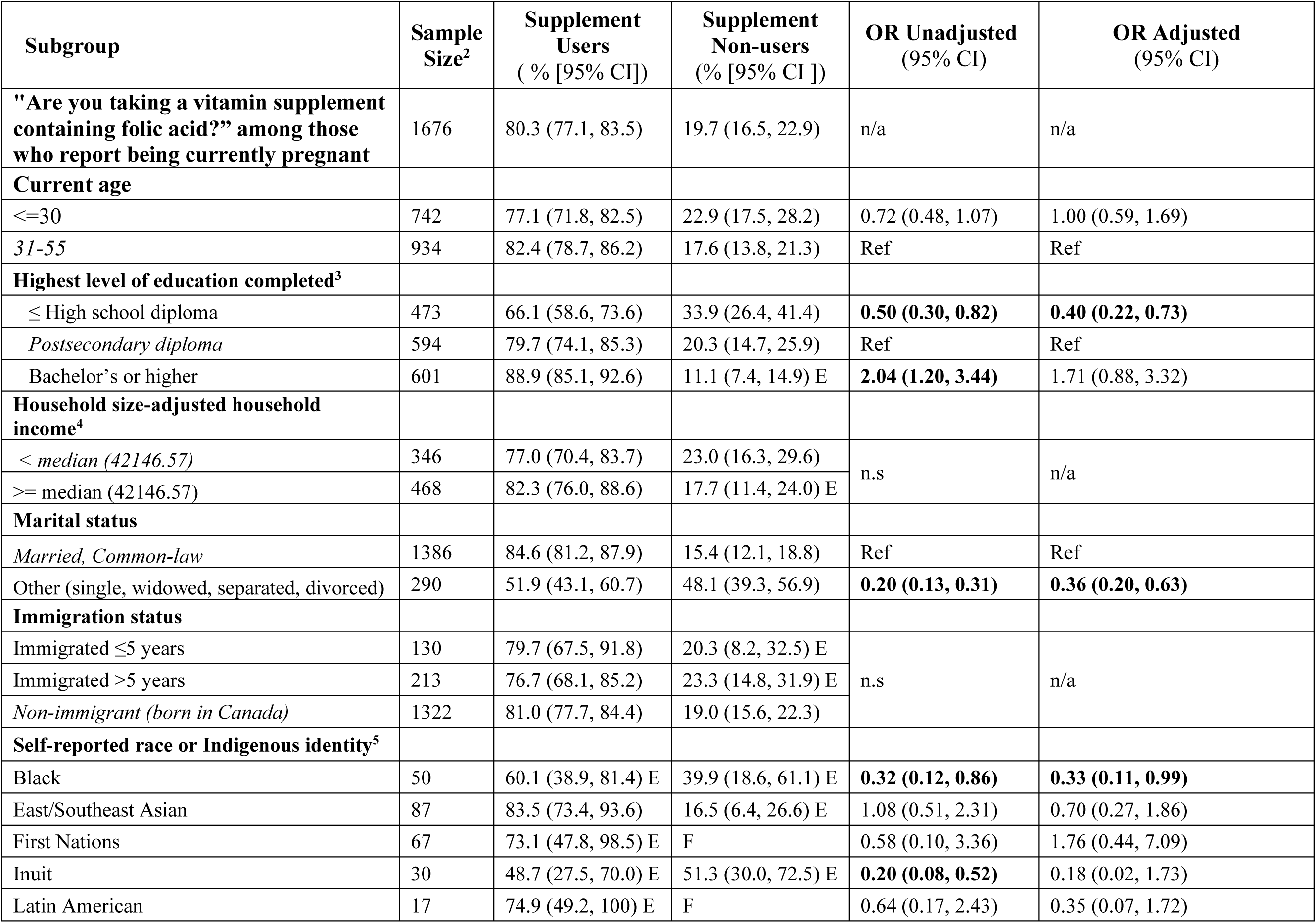
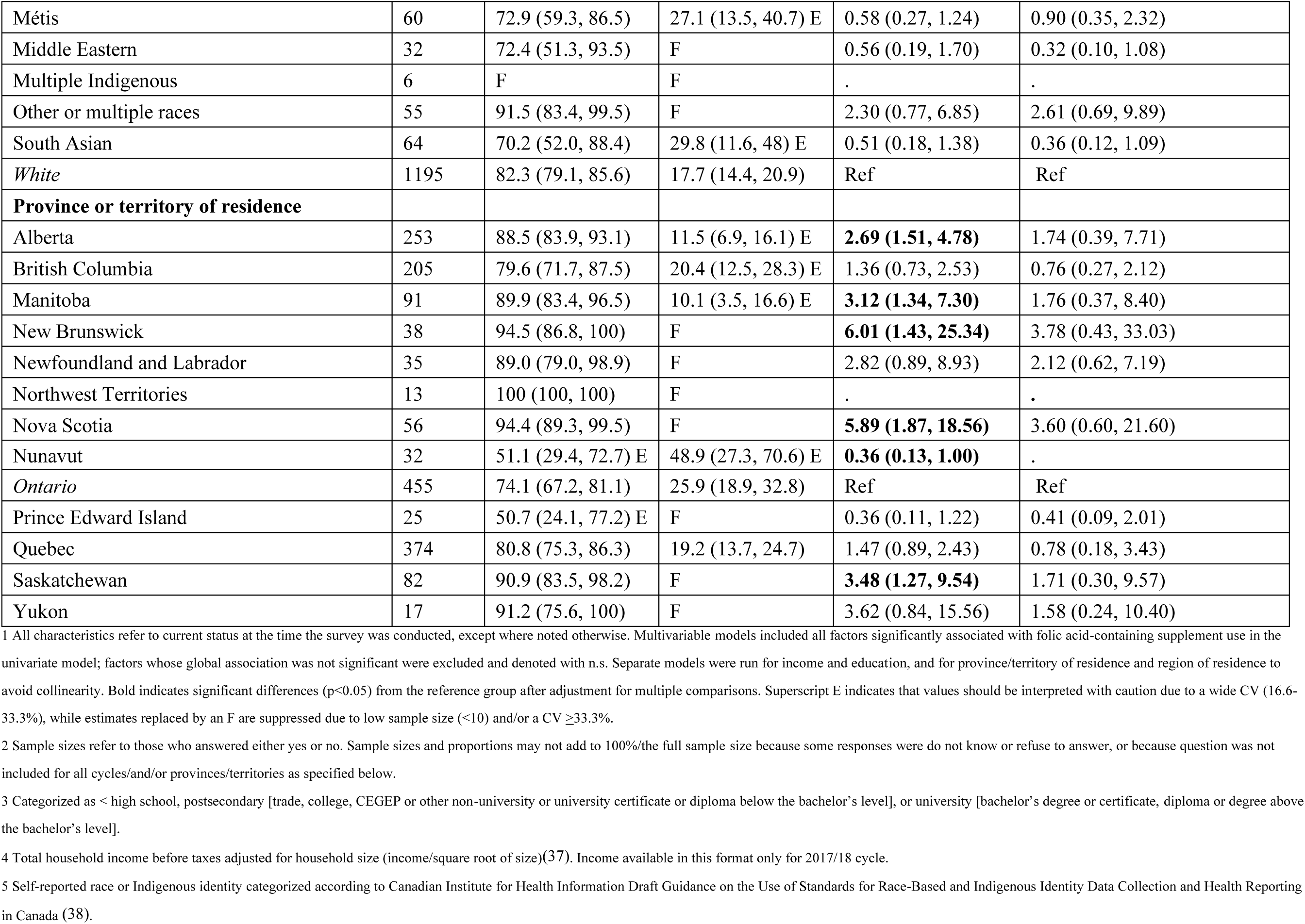
Folic acid-containing supplement use among pregnant females aged 15-55^1^.

**Table 4.**
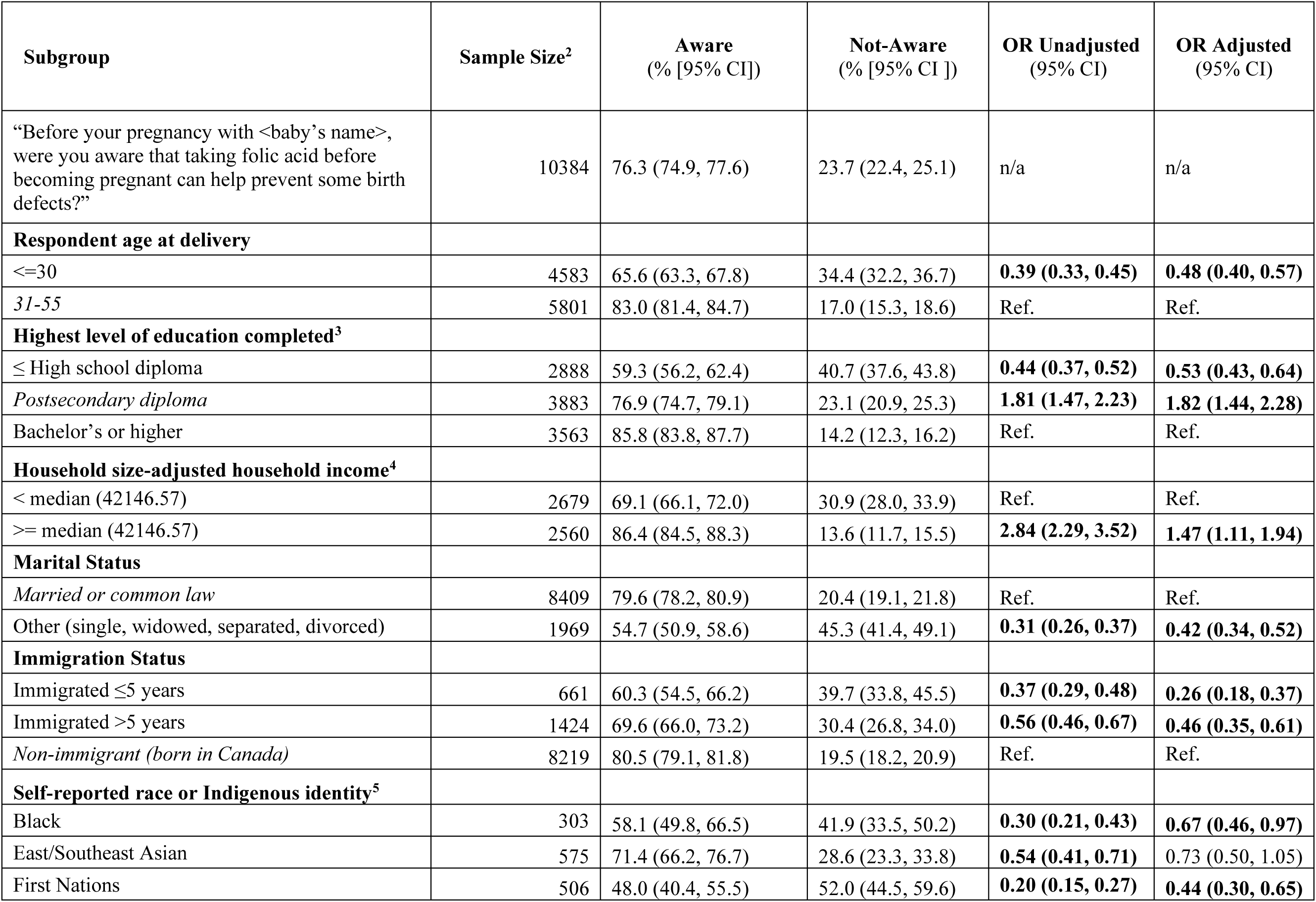
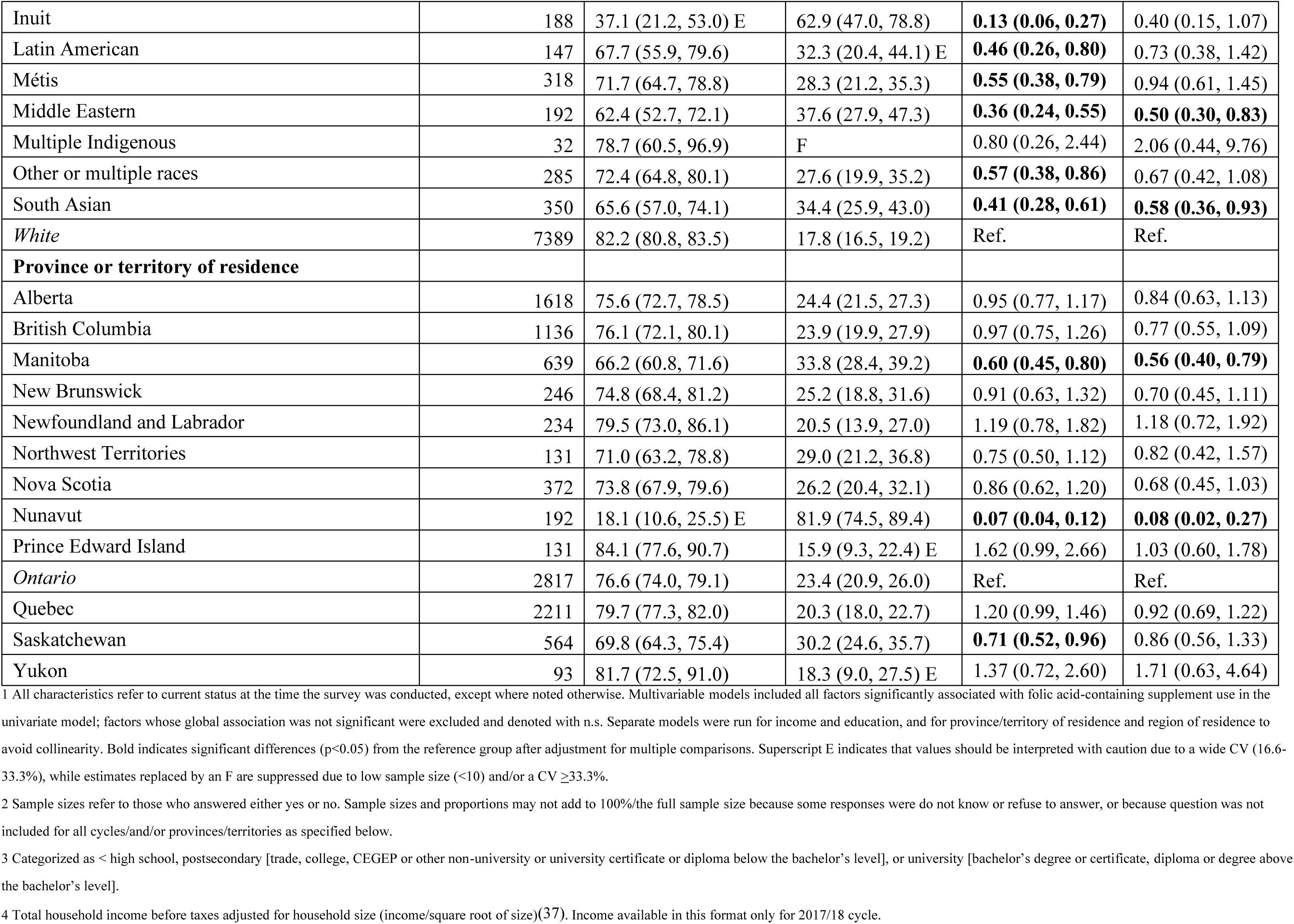
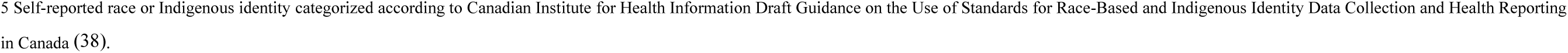
Awareness of link between folic acid and some birth defects^1^.

| Subgroup | Sample Size<sup>2</sup> | Aware<br>(% [95% CI]) | Not-Aware<br>(% [95% CI ]) | OR Unadjusted<br>(95% CI) | OR Adjusted<br>(95% CI) |
| --- | --- | --- | --- | --- | --- |
| “Before your pregnancy with <baby’s name>, were you aware that taking folic acid before becoming pregnant can help prevent some birth defects?” | 10384 | 76.3 (74.9, 77.6) | 23.7 (22.4, 25.1) | n/a | n/a |
| **Respondent age at delivery** |  |  |  |  |  |
| <=30 | 4583 | 65.6 (63.3, 67.8) | 34.4 (32.2, 36.7) | **0.39 (0.33, 0.45)** | **0.48 (0.40, 0.57)** |
| 31-55 | 5801 | 83.0 (81.4, 84.7) | 17.0 (15.3, 18.6) | Ref. | Ref. |
| **Highest level of education completed<sup>3</sup>** |  |  |  |  |  |
| ≤ High school diploma | 2888 | 59.3 (56.2, 62.4) | 40.7 (37.6, 43.8) | **0.44 (0.37, 0.52)** | **0.53 (0.43, 0.64)** |
| Postsecondary diploma | 3883 | 76.9 (74.7, 79.1) | 23.1 (20.9, 25.3) | **1.81 (1.47, 2.23)** | **1.82 (1.44, 2.28)** |
| Bachelor’s or higher | 3563 | 85.8 (83.8, 87.7) | 14.2 (12.3, 16.2) | Ref. | Ref. |
| **Household size-adjusted household income<sup>4</sup>** |  |  |  |  |  |
| < median (42146.57) | 2679 | 69.1 (66.1, 72.0) | 30.9 (28.0, 33.9) | Ref. | Ref. |
| >= median (42146.57) | 2560 | 86.4 (84.5, 88.3) | 13.6 (11.7, 15.5) | **2.84 (2.29, 3.52)** | **1.47 (1.11, 1.94)** |
| **Marital Status** |  |  |  |  |  |
| Married or common law | 8409 | 79.6 (78.2, 80.9) | 20.4 (19.1, 21.8) | Ref. | Ref. |
| Other (single, widowed, separated, divorced) | 1969 | 54.7 (50.9, 58.6) | 45.3 (41.4, 49.1) | **0.31 (0.26, 0.37)** | **0.42 (0.34, 0.52)** |
| **Immigration Status** |  |  |  |  |  |
| Immigrated ≤5 years | 661 | 60.3 (54.5, 66.2) | 39.7 (33.8, 45.5) | **0.37 (0.29, 0.48)** | **0.26 (0.18, 0.37)** |
| Immigrated >5 years | 1424 | 69.6 (66.0, 73.2) | 30.4 (26.8, 34.0) | **0.56 (0.46, 0.67)** | **0.46 (0.35, 0.61)** |
| Non-immigrant (born in Canada) | 8219 | 80.5 (79.1, 81.8) | 19.5 (18.2, 20.9) | Ref. | Ref. |
| **Self-reported race or Indigenous identity<sup>5</sup>** |  |  |  |  |  |
| Black | 303 | 58.1 (49.8, 66.5) | 41.9 (33.5, 50.2) | **0.30 (0.21, 0.43)** | **0.67 (0.46, 0.97)** |
| East/Southeast Asian | 575 | 71.4 (66.2, 76.7) | 28.6 (23.3, 33.8) | **0.54 (0.41, 0.71)** | 0.73 (0.50, 1.05) |
| First Nations | 506 | 48.0 (40.4, 55.5) | 52.0 (44.5, 59.6) | **0.20 (0.15, 0.27)** | **0.44 (0.30, 0.65)** |
| Inuit | 188 | 37.1 (21.2, 53.0) E | 62.9 (47.0, 78.8) | **0.13 (0.06, 0.27)** | 0.40 (0.15, 1.07) |
| Latin American | 147 | 67.7 (55.9, 79.6) | 32.3 (20.4, 44.1) E | **0.46 (0.26, 0.80)** | 0.73 (0.38, 1.42) |
| Métis | 318 | 71.7 (64.7, 78.8) | 28.3 (21.2, 35.3) | **0.55 (0.38, 0.79)** | 0.94 (0.61, 1.45) |
| Middle Eastern | 192 | 62.4 (52.7, 72.1) | 37.6 (27.9, 47.3) | **0.36 (0.24, 0.55)** | **0.50 (0.30, 0.83)** |
| Multiple Indigenous | 32 | 78.7 (60.5, 96.9) | F | 0.80 (0.26, 2.44) | 2.06 (0.44, 9.76) |
| Other or multiple races | 285 | 72.4 (64.8, 80.1) | 27.6 (19.9, 35.2) | **0.57 (0.38, 0.86)** | 0.67 (0.42, 1.08) |
| South Asian | 350 | 65.6 (57.0, 74.1) | 34.4 (25.9, 43.0) | **0.41 (0.28, 0.61)** | **0.58 (0.36, 0.93)** |
| *White* | 7389 | 82.2 (80.8, 83.5) | 17.8 (16.5, 19.2) | Ref. | Ref. |
| **Province or territory of residence** |  |  |  |  |  |
| Alberta | 1618 | 75.6 (72.7, 78.5) | 24.4 (21.5, 27.3) | 0.95 (0.77, 1.17) | 0.84 (0.63, 1.13) |
| British Columbia | 1136 | 76.1 (72.1, 80.1) | 23.9 (19.9, 27.9) | 0.97 (0.75, 1.26) | 0.77 (0.55, 1.09) |
| Manitoba | 639 | 66.2 (60.8, 71.6) | 33.8 (28.4, 39.2) | **0.60 (0.45, 0.80)** | **0.56 (0.40, 0.79)** |
| New Brunswick | 246 | 74.8 (68.4, 81.2) | 25.2 (18.8, 31.6) | 0.91 (0.63, 1.32) | 0.70 (0.45, 1.11) |
| Newfoundland and Labrador | 234 | 79.5 (73.0, 86.1) | 20.5 (13.9, 27.0) | 1.19 (0.78, 1.82) | 1.18 (0.72, 1.92) |
| Northwest Territories | 131 | 71.0 (63.2, 78.8) | 29.0 (21.2, 36.8) | 0.75 (0.50, 1.12) | 0.82 (0.42, 1.57) |
| Nova Scotia | 372 | 73.8 (67.9, 79.6) | 26.2 (20.4, 32.1) | 0.86 (0.62, 1.20) | 0.68 (0.45, 1.03) |
| Nunavut | 192 | 18.1 (10.6, 25.5) E | 81.9 (74.5, 89.4) | **0.07 (0.04, 0.12)** | **0.08 (0.02, 0.27)** |
| Prince Edward Island | 131 | 84.1 (77.6, 90.7) | 15.9 (9.3, 22.4) E | 1.62 (0.99, 2.66) | 1.03 (0.60, 1.78) |
| *Ontario* | 2817 | 76.6 (74.0, 79.1) | 23.4 (20.9, 26.0) | Ref. | Ref. |
| Quebec | 2211 | 79.7 (77.3, 82.0) | 20.3 (18.0, 22.7) | 1.20 (0.99, 1.46) | 0.92 (0.69, 1.22) |
| Saskatchewan | 564 | 69.8 (64.3, 75.4) | 30.2 (24.6, 35.7) | **0.71 (0.52, 0.96)** | 0.86 (0.56, 1.33) |
| Yukon | 93 | 81.7 (72.5, 91.0) | 18.3 (9.0, 27.5) E | 1.37 (0.72, 2.60) | 1.71 (0.63, 4.64) |

1 All characteristics refer to current status at the time the survey was conducted, except where noted otherwise. Multivariable models included all factors significantly associated with folic acid-containing supplement use in the univariate model; factors whose global association was not significant were excluded and denoted with n.s. Separate models were run for income and education, and for province/territory of residence and region of residence to avoid collinearity. Bold indicates significant differences ( $p < 0.05$ ) from the reference group after adjustment for multiple comparisons. Superscript E indicates that values should be interpreted with caution due to a wide CV (16.6-33.3%), while estimates replaced by an F are suppressed due to low sample size ( $< 10$ ) and/or a CV $\geq 33.3\%$ .
2 Sample sizes refer to those who answered either yes or no. Sample sizes and proportions may not add to 100%/the full sample size because some responses were do not know or refuse to answer, or because question was not included for all cycles/and/or provinces/territories as specified below.
3 Categorized as < high school, postsecondary [trade, college, CEGEP or other non-university or university certificate or diploma below the bachelor's level], or university [bachelor's degree or certificate, diploma or degree above the bachelor's level].
4 Total household income before taxes adjusted for household size (income/square root of size)(37). Income available in this format only for 2017/18 cycle.
5 Self-reported race or Indigenous identity categorized according to Canadian Institute for Health Information Draft Guidance on the Use of Standards for Race-Based and Indigenous Identity Data Collection and Health Reporting in Canada (38).

**Table 5.** Folic acid-containing supplement in the 3 months prior to becoming pregnant^1^.

| Subgroup | Sample Size<sup>2</sup> | Supplement Users<br>(% [95% CI]) | Supplement Non-users<br>(% [95% CI ]) | OR Unadjusted<br>(95% CI) | OR Adjusted<br>(95% CI) |
| --- | --- | --- | --- | --- | --- |
| “In the three months before you got pregnant with <baby’s name>, did you take a folic acid supplement or a multivitamin containing folic acid?” | 10326 | 63.7 (62.2-65.1) | 36.3 (34.9-37.8) | n/a | N/a |
| Frequency of supplement use among those who used supplements |  |  |  |  |  |
| Every day or almost every day | 6201 | 98.3 (97.9, 98.8) | n/a | n/a | n/a |
| Less than every day or almost every day | 106 | 1.6 (1.1, 2.0) |  |  |  |
| Aware prior to pregnancy of link between folic acid and birth defects |  |  |  |  |  |
| Yes | 7731 | 74.0 (72.4, 75.6) | 26.0 (24.4, 27.6) | Ref | Ref |
| No | 2564 | 30.1 (27.1, 33.2) | 69.9 (66.8, 72.9) | **0.15 (0.13, 0.18)** | **0.20 (0.17, 0.24)** |
| Respondent age at delivery |  |  |  |  |  |
| <=30 | 4557 | 53.8 (51.6, 56.0) | 46.2 (44.0, 48.4) | **0.50 (0.44, 0.57)** | **0.80 (0.69, 0.93)** |
| 31-55 | 5769 | 69.9 (68.0, 71.8) | 30.1 (28.2, 32.0) | Ref | Ref |
| Highest level of education completed<sup>3</sup> |  |  |  |  |  |
| ≤ High school diploma | 2859 | 44.8 (41.9, 47.6) | 55.2 (52.4, 58.1) | **0.46 (0.39, 0.53)** | **0.64 (0.53, 0.76)** |
| Postsecondary diploma | 3868 | 63.9 (61.5, 66.4) | 36.1 (33.6, 38.5) | Ref | Ref |
| Bachelor’s or higher | 3549 | 74.3 (72.0, 76.6) | 25.7 (23.4, 28.0) | **1.63 (1.39, 1.92)** | **1.27 (1.08, 1.51)** |
| Household size-adjusted household income<sup>4</sup> |  |  |  |  |  |
| < median (42146.57) | 2678 | 53.4 (50.2, 56.5) | 46.6 (43.5, 49.8) | Ref | Ref |
| >= median (42146.57) | 2541 | 73.7 (70.9, 76.5) | 26.3 (23.5, 29.1) | **2.45 (2.01, 2.98)** | **1.74 (1.38, 2.21)** |
| Marital status |  |  |  |  |  |
| *Married or common law* | 8359 | 67.4 (65.8, 69.0) | 32.6 (31.0, 34.2) | Ref | Ref |
| Other (single, widowed, separated, divorced) | 1961 | 39.3 (35.8, 42.9) | 60.7 (57.1, 64.2) | **0.31 (0.27, 0.37)** | **0.52 (0.43, 0.64)** |
| **Immigration status** |  |  |  |  |  |
| Immigrated ≤5 years | 656 | 57.5 (51.1, 63.8) | 42.5 (36.2, 48.9) | **0.74 (0.56, 0.96)** | 0.78 (0.54, 1.15) |
| Immigrated >5 years | 1421 | 63.5 (59.5, 67.5) | 36.5 (32.5, 40.5) | 0.95 (0.79, 1.15) | 0.93 (0.69, 1.27) |
| *Non-immigrant (born in Canada)* | 8169 | 64.7 (63.1, 66.2) | 35.3 (33.8, 36.9) | Ref | Ref |
| **Self-reported race or Indigenous identity<sup>5</sup>** |  |  |  |  |  |
| Black | 303 | 52.0 (43.7, 60.3) | 48.0 (39.7, 56.3) | **0.53 (0.38, 0.75)** | 0.95 (0.56, 1.61) |
| East/Southeast Asian | 575 | 68.0 (63.1, 72.9) | 32.0 (27.1, 36.9) | 1.04 (0.82, 1.32) | 1.14 (0.79, 1.65) |
| First Nations | 502 | 31.3 (25.5, 37.2) | 68.7 (62.8, 74.5) | **0.22 (0.17, 0.30)** | **0.54 (0.39, 0.75)** |
| Inuit | 184 | 34.0 (17.6, 50.3) E | 66.0 (49.7, 82.4) | **0.25 (0.11, 0.56)** | 0.99 (0.36, 2.70) |
| Latin American | 146 | 66.9 (54.9, 79.0) | 33.1 (21.0, 45.1) E | 0.99 (0.57, 1.73) | 1.37 (0.65, 2.89) |
| Métis | 315 | 47.2 (38.1, 56.3) | 52.8 (43.7, 61.9) | **0.44 (0.30, 0.64)** | 0.65 (0.40, 1.07) |
| Middle Eastern | 191 | 60.2 (49.9, 70.5) | 39.8 (29.5, 50.1) | 0.74 (0.48, 1.15) | 0.97 (0.57, 1.64) |
| Multiple Indigenous | 32 | 57.9 (32.7, 83.2) E | 42.1 (16.8, 67.3) E | 0.68 (0.22, 2.09) | 1.37 (0.30, 6.20) |
| Other or multiple races | 278 | 62.1 (53.5, 70.7) | 37.9 (29.3, 46.5) | 0.81 (0.56, 1.17) | 0.85 (0.57, 1.27) |
| South Asian | 349 | 58.1 (49.6, 66.5) | 41.9 (33.5, 50.4) | **0.68 (0.47, 0.97)** | 0.80 (0.52, 1.23) |
| *White* | 7352 | 67.1 (65.5, 68.7) | 32.9 (31.3, 34.5) | Ref | Ref |
| **Province or territory of residence** |  |  |  |  |  |
| Alberta | 1609 | 60.5 (57.3, 63.6) | 39.5 (36.4, 42.7) | 0.86 (0.71, 1.02) | 0.88 (0.67, 1.16) |
| British Columbia | 1129 | 66.5 (62.2, 70.8) | 33.5 (29.2, 37.8) | 1.11 (0.88, 1.40) | 1.10 (0.82, 1.47) |
| Manitoba | 634 | 55.4 (49.7, 61.0) | 44.6 (39.0, 50.3) | **0.69 (0.54, 0.89)** | 0.87 (0.64, 1.20) |
| New Brunswick | 246 | 57.8 (50.2, 65.3) | 42.2 (34.7, 49.8) | 0.76 (0.55, 1.06) | 0.84 (0.55, 1.28) |
| Newfoundland and Labrador | 234 | 49.1 (40.4, 57.8) | 50.9 (42.2, 59.6) | **0.54 (0.37, 0.78)** | **0.55 (0.35, 0.87)** |
| Northwest Territories | 127 | 54.4 (42.9, 65.8) | 45.6 (34.2, 57.1) | 0.67 (0.41, 1.08) | 0.92 (0.45, 1.86) |
| Nova Scotia | 370 | 53.9 (47.0, 60.8) | 46.1 (39.2, 53.0) | **0.65 (0.48, 0.88)** | 0.72 (0.49, 1.06) |
| Nunavut | 190 | 22.4 (13.8, 30.9) E | 77.6 (69.1, 86.2) | **0.16 (0.10, 0.27)** | 0.52 (0.12, 2.31) |
| Prince Edward Island | 131 | 68.1 (59.0, 77.3) | 31.9 (22.7, 41.0) | 1.20 (0.77, 1.85) | 1.09 (0.65, 1.85) |
| *Ontario* | 2802 | 64.2 (61.3, 67.0) | 35.8 (33.0, 38.7) | Ref | Ref |
| Quebec | 2198 | 68.5 (65.7, 71.2) | 31.5 (28.8, 34.3) | **1.21 (1.02, 1.45)** | 1.18 (0.91, 1.55) |
| Saskatchewan | 563 | 57.5 (51.7, 63.3) | 42.5 (36.7, 48.3) | **0.76 (0.58, 0.99)** | 1.10 (0.76, 1.60) |
| Yukon | 93 | 55.6 (43.5, 67.7) | 44.4 (32.3, 56.5) | 0.70 (0.42, 1.17) | 0.63 (0.36, 1.09) |

1 All characteristics refer to current status at the time the survey was conducted, except where noted otherwise. Multivariable models included all factors significantly associated with folic acid-containing supplement use in the univariate model; factors whose global association was not significant were excluded and denoted with n.s. Separate models were run for income and education, and for province/territory of residence and region of residence to avoid collinearity. Bold indicates significant differences ( $p < 0.05$ ) from the reference group after adjustment for multiple comparisons. Superscript E indicates that values should be interpreted with caution due to a wide CV (16.6-33.3%), while estimates replaced by an F are suppressed due to low sample size ( $< 10$ ) and/or a CV $\geq 33.3\%$ .
2 Sample sizes refer to those who answered either yes or no. Proportions may not add to 100%/the full sample size because some responses were do not know or refuse to answer, or because question was not included for all cycles/and/or provinces/territories as specified below.
3 Categorized as < high school, postsecondary [trade, college, CEGEP or other non-university or university certificate or diploma below the bachelor's level], or university [bachelor's degree or certificate, diploma or degree above the bachelor's level].
4 Total household income before taxes adjusted for household size (income/square root of size)(37). Income available in this format only for 2017/18 cycle.
5 Self-reported race or Indigenous identity categorized according to Canadian Institute for Health Information Draft Guidance on the Use of Standards for Race-Based and Indigenous Identity Data Collection and Health Reporting in Canada(38).

**Table 6.**
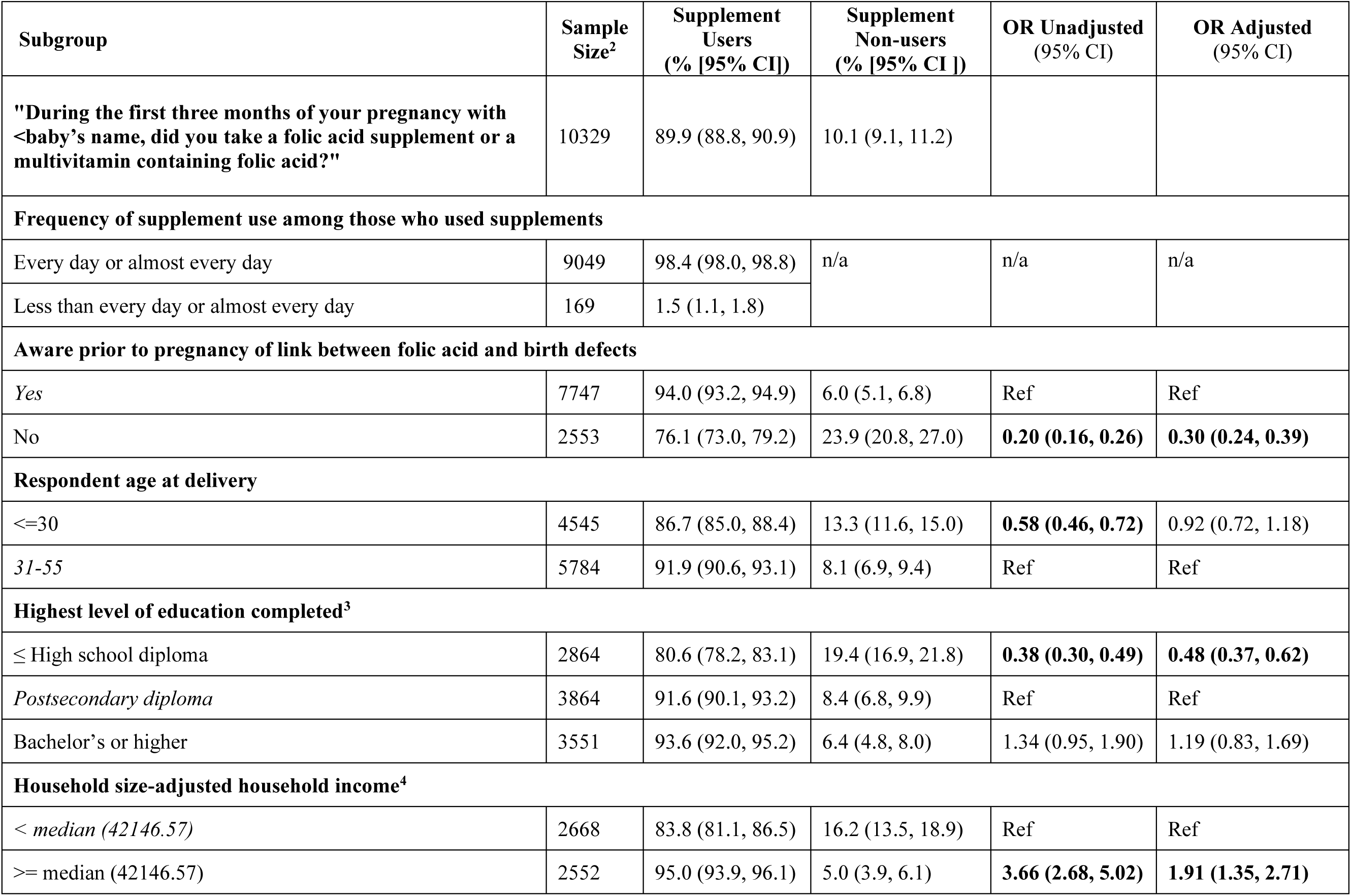
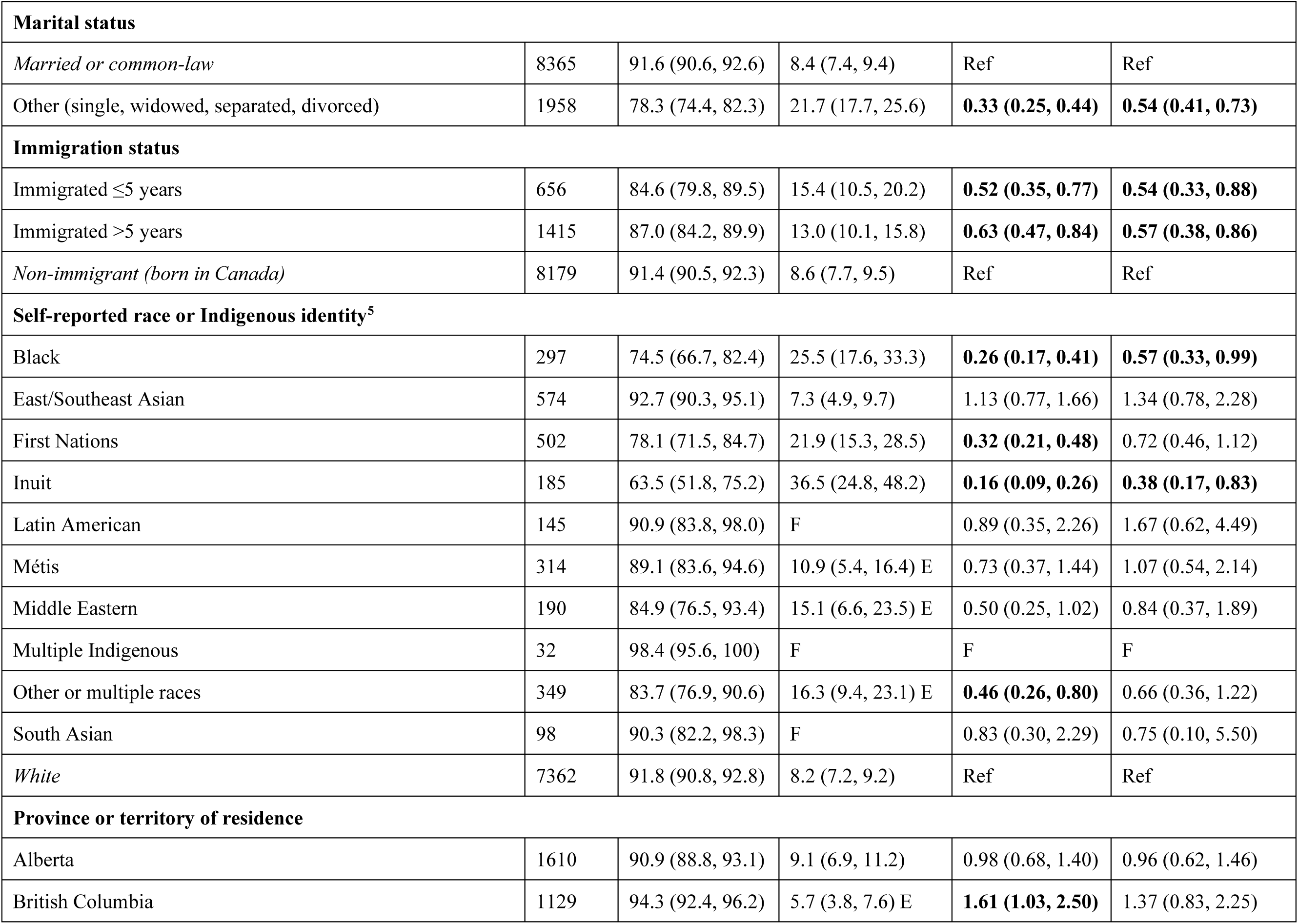
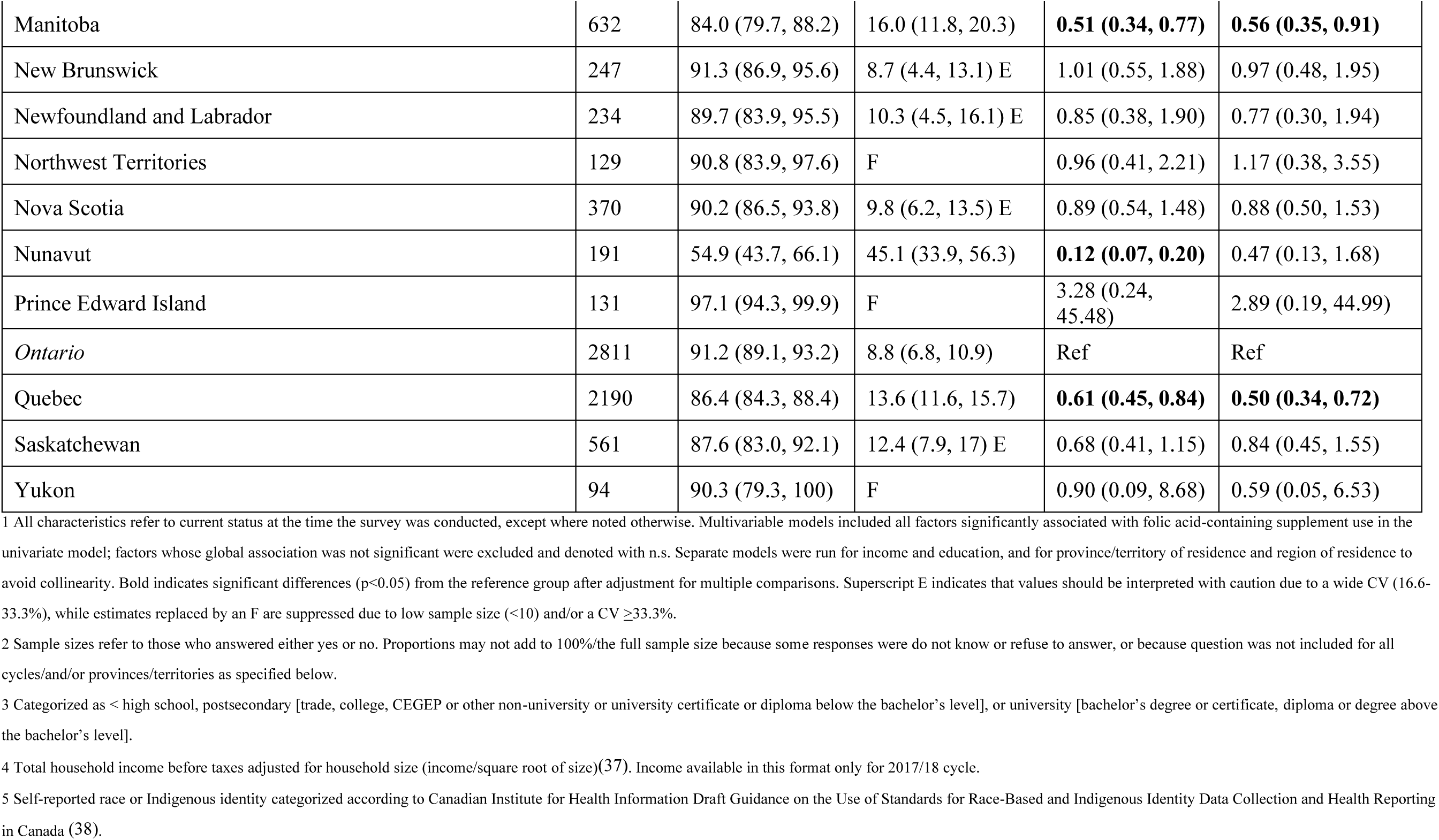
Folic acid-containing supplement use in the first three months of pregnancy^1^.

### Factors associated with folic acid-containing supplement use

For ease of interpretation, heat maps summarizing the odds ratios for the associations between folic acid-containing supplement use and the sociodemographic, geographic, and health factors examined are shown in **Figures 2A** (unadjusted) and **2B** (adjusted for significant factors from univariate models, see Tables 1-6 and Supplemental Tables 1-6 for details), with detailed results shown in the relevant tables. Data for some characteristics (source of household income, region of residence, type of community, BMI category, food security, consumption of green vegetables, consumption of total fruits and vegetables, frequency of general physical check-up, alcohol use, smoking, and years since birth of most recent infant [for those who reported giving birth in the past five years only]) are shown in the corresponding **Supplemental Tables 1-6**. In the interest of brevity and to decrease reliance on p-values, we focus our descriptions on patterns of associations that were consistent between similar outcomes and between the unadjusted and adjusted models. Unless otherwise stated, ORs provided in the text are for the adjusted model.

**Figure 2.**
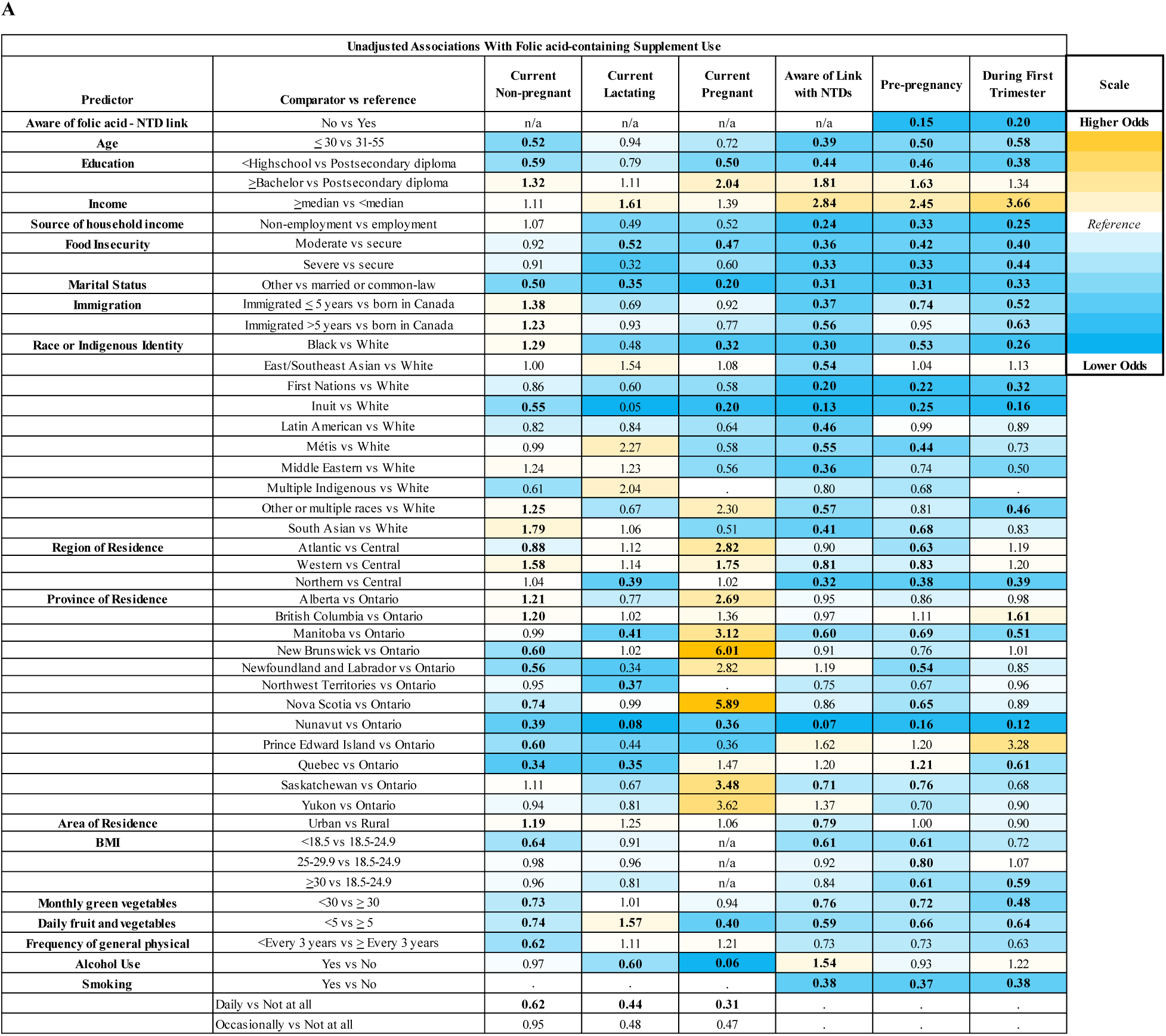
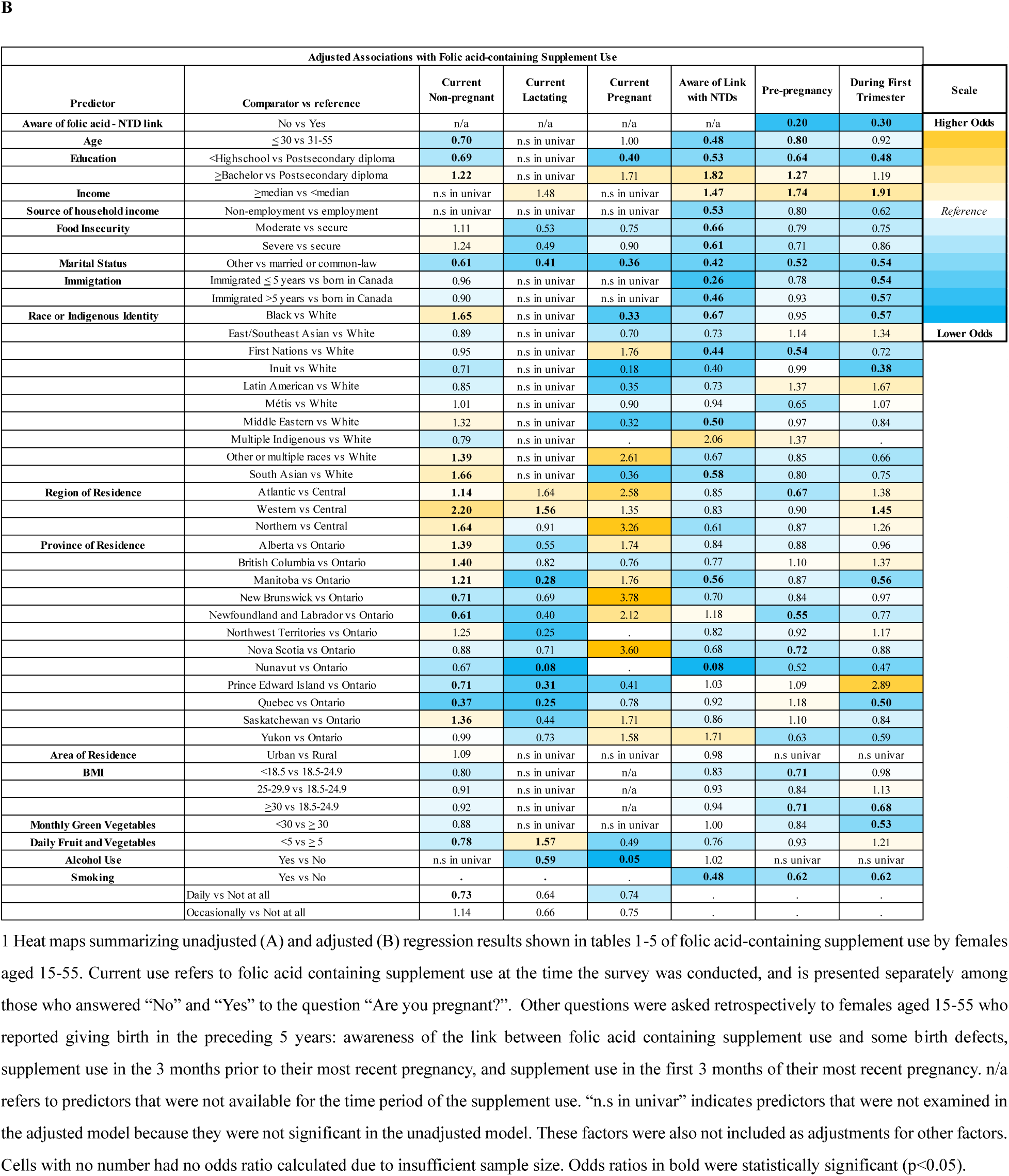
Summary of regression results^1^.

Among respondents who had given birth in the preceding 5 years, awareness of the link between folic acid-containing supplements and some birth defects was the strongest predictor of supplement use. Those who were unaware of the link pre-pregnancy were less likely to report using folic acid-containing supplements in the 3 months prior to pregnancy (OR 0.20 [95% CI: 0.17, 0.24]) and in the first 3 months of pregnancy (OR 0.30 [95% CI: 0.24, 0.39]).

Younger age was associated with a lower likelihood of using a folic acid-containing supplement currently among non-pregnant respondents (0.70 [95% CI: 0.63, 0.79]), and retrospectively among people who had given birth in the preceding 5 years pre-pregnancy (OR 0.80 [95% CI: 0.69, 0.93]), as well as with a lower odds of being aware of the link between folic acid and some birth defects (OR 0.48 [95% CI: 0.40, 0.57]). A similar trend was observed for folic acid-containing supplement use during pregnancy among those who had given birth in the last 5 years, though this did not remain significant upon adjustment.

Educational attainment was associated with supplement use among non-pregnant and pregnant women, and awareness of the link between folic acid and some birth defects, with adjusted odds ratios ranging from 0.4 to 0.7 for high school compared to postsecondary, and 1.2 to 1.8 for bachelor’s degree or higher compared to postsecondary diploma. Marital status was associated with folic acid-containing supplement use at all outcomes, with respondents who reported not being married or in a common law relationship less likely to report taking a folic acid-containing supplement (ORs from 0.4 to 0.6) or to be aware of the link between folic acid and some birth defects (OR 0.42 [95% CI: 0.34, 0.52]).

Having a household income ≥median was associated with a higher odds of taking a folic acid-containing supplement peri-conceptionally (OR 1.74 [95% CI: 1.38, 2.21] and 1.91 [95% CI: 1.35, 2.71] for the three months prior to and first 3 months of pregnancy respectively) and with a higher odds of being aware of the link between folic acid and some birth defects (OR 1.47 [95% CI: 1.11, 1.94]) among respondents who had given birth in the preceding 5 years. A similar trend was observed in the unadjusted model for current supplement use among pregnant, lactating, and non-pregnant respondents, but this did not reach statistical significance. Main source of household income from non-employment sources (social assistance, welfare, alimony or other) tended to associate with lower odds of supplement use during all life-stages, but this was significant in the adjusted model only for awareness of the link between folic acid and some birth defects (OR 0.53 [95% CI: 0.34, 0.82]). Moderate or severe food insecurity was consistently associated with lower odds of all outcomes, though this remained significant in the adjusted model only for awareness of the link between folic acid and some birth defects (ORs 0.61-0.66).

Self-reported race or Indigenous identity demonstrated variable associations with folic acid-containing supplement use. For example, in the adjusted model, self-reported Black or South Asian race was associated with a higher odds of current folic acid-containing supplement use than those reporting a white race among non-pregnant respondents (OR 1.65 [95% CI: 1.28, 2.12]) and 1.66 [95% CI: 1.32, 2.08] respectively), but tended to associate with lower odds of folic acid-containing supplement use during pregnancy, though this was not always significant. There was a pattern of lower odds of folic-acid containing supplement use in the 3 months before and first 3 months of pregnancy among respondents who self-reported a non-white race or Indigenous identity relative to those reporting a white race, though upon adjustment, this remained significant only for the first 3 months of pregnancy among those who identified as Black or Inuit (OR 0.57 [95% CI: 0.33, 0.99] and 0.38 [95% CI: 0.17, 0.83] respectively), and only for the 3 months prior to their most recent pregnancy among those who identified as First Nations (OR 0.54 [95% CI: 0.39, 0.75]). All racial groups or Indigenous identities had a lower odds of reporting awareness of the link between folic acid and some birth defects relative to those reporting white race in the unadjusted mode, though in the adjusted model this remained significant only for respondents who identified as Black (OR 0.67 [95% CI: 0.46, 0.97]), Middle Eastern (OR 0.50 [95% CI: 0.30, 0.83]), South Asian (OR 0.58 [95% CI: 0.36, 0.93]) or First Nations (OR 0.44 [95% CI: 0.30, 0.65]).

Among respondents who had given birth in the preceding 5 years, both recent and established immigrants to Canada had a lower odds of being aware of the link between folic acid and some birth defects (ORs 0.3-0.5), and were less likely to report having used a folic acid-containing supplement in the first 3 months of pregnancy (ORs 0.5-0.6) compared to those born in Canada. We identified no consistent pattern for associations with folic acid-containing supplement use by immigration among non-pregnant or lactating respondents, or those who reported being pregnant at the time of the survey.

Associations by region or province of residence differed between outcomes, and few associations remained significant upon adjustment for other factors. Though this was not always statistically significant, Nunavut consistently had a lower odds of folic acid-containing supplement use relative to Ontario across life stages examined, and a substantially lower odds of reporting awareness of a link between folic acid and some birth defects (OR 0.08 (95% CI: 0.02, 0.27).

BMI in the underweight or obese range was associated with a lower odds of folic acid-containing supplement use relative to those in the normal range in the 3 months prior to pregnancy among those who had given birth in the preceding 5 years (OR 0.71 for both), while BMI in the obese range was additionally associated with a lower odds of supplement use in the first three months of pregnancy (OR: 0.68 [95% CI: 0.46, <1.00]). BMI in the overweight range was not significantly associated with supplement use in any of the life stages examined. Likewise, BMI was not associated with awareness of the link between folic acid and birth defects in the adjusted model.

Monthly intake of green vegetables or daily intake of fruits and vegetables was generally associated with lower odds of supplement use among non-pregnant and pregnant respondents, as well as during the periconceptional period in the unadjusted model, but this was attenuated upon adjustment. In contrast, for respondents who reported currently lactating, a lower intake of daily fruits and vegetables was associated with a higher odds of using a folic acid-containing supplement (OR 1.57 [1.01, 2.45]). Similarly, smoking consistently showed strong negative associations with folic acid supplement use or awareness of the link between folic acid and some birth defects in the unadjusted model (ORs 0.3-0.6), but was no longer significant upon adjustment for current supplement use among respondents who were lactating or pregnant. Alcohol use was negatively associated with folic acid-containing supplement use during current pregnancy and lactation (ORs 0.05-0.59) but was not associated with supplement use among non-pregnant women of childbearing age, or with awareness of the link with NTDs in the adjusted model. Frequency of general physical checkup was only available in a small subset of respondents for the 2015/16 survey, and was associated only with supplement use among non-pregnant women in the unadjusted model (OR 0.62 [95% CI: 0.44, 0.87]).

Other studies have identified pregnancy intention as a determinant of folic acid-containing supplement use (22), but this variable was not available within our dataset. In an exploratory analysis, we examined folic acid-containing supplement use among non-pregnant females of childbearing age stratified by reported sexual behaviors as a proxy. Respondent who were not sexually active with males or who reported using a form of birth control were significantly less likely to report using a folic acid-containing supplement (OR 0.24, [95% CI: 0.20, 0.30] and 0.63 [0.56, 0.70] respectively, data not shown) than those who reported being sexually active with males and who did not use a form of birth control.

## DISCUSSION

Our analysis provides recent, nationally representative estimates of folic acid-containing supplement use among females of reproductive age in Canada. These data are highly policy-relevant given Canadian recommendations that anyone who could become pregnant consume a daily multivitamin supplement containing folic acid, whether they are planning a pregnancy or not, continuing throughout pregnancy and lactation. Our paper provides data about supplement use pre-pregnancy, during the first trimester, and among females who are non-pregnant, pregnant, and, to our knowledge for the first time in national Canadian data, lactating. Previous nationally representative data from the Maternal Experiences Survey (MES) is now >15 years old and included only retrospective data on the 3 months before and first 3 months of pregnancy (23). A more recent analysis of the CCHS Maternal Experiences module investigated only awareness of the link between folic acid and some birth defects, and did not include disaggregation by race or Indigenous identity (24). Analyses of data from the CCHS Nutrition-2015 excluded pregnant and lactating women (25). Other Canadian research cohorts studies are not representative of the Canadian population, cannot provide provincial or territorial estimates, and are considerably smaller than the present analysis, which includes over 50 thousand respondents weighted to represent over 9 million women living in Canada. Our large, diverse sample also enabled us to look at associations between sociodemographic and health characteristics and supplement use, which are essential for assessing inequalities in Canada. These findings will also be informative for other countries, including the United States, which has similar initiatives.

The proportion of respondents who reported using folic acid in the present analysis is higher than previously reported in the MES for 2006-7 for the 3 months prior to pregnancy (63.7% [95% CI: 62.2-65.1] vs 57.7% [95% CI: 56.4–59.0]), but is similar for the first 3 months of pregnancy (89.9% [95% CI: 88.8, 90.9] vs 89.7% [95% CI: 95% CI: 88.9–90.5]). The proportion of respondents who reported being aware prior to pregnancy that folic acid supplementation could help prevent some birth defects in our study (76.3% [95% CI: 74.9, 77.6]) was essentially unchanged compared to that reported in the 2006-7 MES (77.6% [95% CI: 76.6–78.7]), and was comparable to a recently published analysis using data from the 2017-18 CCHS (79%) (24). The prevalence of folic acid-containing supplement use during pregnancy in our nationally representative sample (80.3% [95% CI: 77.1, 83.5]) is within the range reported in other non-nationally representative Canadian cohorts; for example lower than reported in the Alberta Pregnancy Outcomes and Nutrition cohort (APrON, 91-97% (27)), but higher than reported in Maternal-Infant Research on Environmental Chemicals cohort (MIREC, 78%, (26)).

In our analysis, estimates of folic acid-containing supplement use among non-pregnant females of childbearing age are lower than reported previously from the 2015 CCHS-Nutrition: 11.0% (95% CI: 10.2, 11.8) for those aged ≤30, and 19.2% ( 95% CI: 18.5, 19.9) for those aged 31-55 in our study compared to 15.4% (SE 1.7) of those aged 14-18, 19.1% (SE 2.7) of those aged 19-30, and 24.7% (SE 1.7) of those aged 31-50 (25). This lower prevalence may be explained by differences in how folic acid containing supplement use was defined. The 2015 CCHS-Nutrition analysis classified respondents as supplement users if they reported using a supplement at least once in the previous 30 days in a food frequency questionnaire, whereas in our study, it was assessed by the question “are you taking a vitamin supplement containing folic acid?” which may not have captured sporadic or occasional supplement users.

Canadian public health guidelines recommend that anyone who could become pregnant, is pregnant, or is lactating consume a daily multivitamin supplement containing 400 µg of folic acid (15). Taken together, our data suggest that adherence to Canadian guidelines to consume a supplement with folic acid is low among the general population of non-pregnant females of childbearing age. Despite this, a majority report using a supplement in line with the recommendation during pregnancy, and to a lesser extent before pregnancy and in lactation.

In our analysis, respondents from Nunavut consistently reported lower prevalence of folic acid-containing supplement use across measures compared to respondents from other provinces or territories, as well as a substantially lower prevalence of awareness of the link between folic acid and some birth defects. While the associations were attenuated upon adjustment for other factors, such as age, marital status, income, and food insecurity, the pattern remained consistent. These results are similar to findings from the 2006-7 Maternity Experiences survey (23). Our results are also similar to the proportion of folic acid-containing supplement users among non-pregnant women of reproductive age in the 2007-8 Inuit Health Survey (8.1 vs 6.8%), the majority of whom were from Nunavut (23, 31). The identification of this persistent inequality may be important for developing targeted public health initiatives.

Our study also identified other sociodemographic and health factors associated with folic acid-containing supplement use or awareness of the link between folic acid and some birth defects. Notably, younger age, single marital status, lower educational attainment, income below the median, and smoking tended to associate with lower odds of using a supplement or of being aware of the link between folic acid and some birth defects. Similar associations were identified between general vitamin/mineral supplement use and respondent age, education, income, BMI, and smoking in the 2015 CCHS-Nutrition (25). These factors have also been associated with supplement use in other studies within Canada and the United States (22, 32).

The factors associated with lower odds of folic acid supplement use in our study reflect those associated with lower folate status. In an analysis of red blood cell folate status of female supplement non-users of childbearing age in the Canadian Health Measures Survey, those with younger age, lower educational attainment, lower income, or current smoking were more likely to have low folate status compared to other supplement non-users (33). Similar associations have been reported in U.S cohorts summarized by the Environmental influences on Child Health Outcomes (ECHO) consortium (34). This suggests that these subgroups of supplement non-users remain at higher risk for having an NTD-affected pregnancy despite mandatory folic acid fortification.

Strengths of our study include its large, nationally representative sample, which enabled provincial and territorial-level estimates, the collection of detailed sociodemographic and health data, and the examination of folic acid-containing supplement use at multiple periods relevant both to the prevention of NTDs and to Canadian guidelines. Our study also has several limitations. The questions regarding folic acid-containing supplement use in the CCHS were broad “yes” or “no” questions, without information available on dose. In addition, while our analysis shows that the vast majority (>98%) of those who used folic acid-containing supplements peri-conceptionally reported taking them every day or almost every day, information on frequency of supplement use was unavailable for the question on current folic acid-containing supplement use. Further, questions referred to “folic acid”, a term in line with Canadian public health guidance specifying the use of this form of folate for NTD prevention (35) however, this may have caused an underestimation of supplement use among those unfamiliar with this term, or who used supplements containing another form of folate. There was also no information available in our survey on the trimester of current pregnancy, so it is possible that some of the pregnant respondents may have used supplements in the first trimester, but discontinued use thereafter. Another limitation of the retrospective nature of the questions about folic acid-containing supplement use peri-conceptionally is that many participant characteristics examined reflect the time at which the survey was conducted, which may have changed in the up to 5 years since respondents last gave birth. The associations with many of these characteristics, such as income, and education, were consistent between current and retrospective supplement use, which may give more confidence that these are important determinants despite this limitation.

## CONCLUSION

While the majority of people living in Canada who are or have been pregnant report using folic acid-containing supplements peri-conceptionally, adherence to Canadian guidelines on folic acid supplementation among the general population of non-pregnant females of childbearing age is <20%, and inequalities remain by factors such as age, income, education, marital status, and geographic location. Awareness that folic acid supplements can prevent some birth defects is a strong predictor of supplement use, which makes it an important message for inclusion in public health education initiatives to improve adherence to guidelines.

## Authors Contributions

All authors contributed to the design of the research, development of the analysis plan, study oversight, interpretation of results, and review and approval of the final manuscript. LL and TML performed the statistical analysis. KEH wrote the article with input from all coauthors.

## Data Availability

Data described in the manuscript, code book, and analytic code will be made available upon request pending application and approval to Statistics Canada and Health Canada.

## Editorial Board Membership

Hope Weiler is an Editorial Board Member for The Journal of Nutrition and played no role in the Journal’s evaluation of the manuscript.

## Abbreviations

BMI: Body Mass Index
CCHS: Canadian Community Health Survey
CI: Confidence Interval
CV: Coefficient of Variation
NHANES: National Health and Nutrition Examination Survey
NTD: Neural Tube Defects

## Clinical trial registry number

not applicable

## Registry and registry number for systematic reviews of meta-analysis

not applicable

