## Supplemental Material for "Folic acid-containing supplement use among females aged 15-55 in the Canadian Community Health Survey 2015-2018"

### Supplemental Material – Table of Contents

**Supplemental Materials and Methods**

*Folic acid-containing supplement use*

Surveys were administered by a trained interviewer. Current supplement use was obtained from the question “Are you taking a vitamin supplement containing folic acid?”. Interviewers had access to a glossary of terms that included a definition for folic acid (“A vitamin of the B complex, used before and during pregnancy to prevent some birth defects”), and were instructed to select the “Yes” if the respondent took a prenatal vitamin but was unsure what it contained. This was because a “prenatal” multivitamin in Canada must contain folate, which for the vast majority of products is folic acid (1). Pregnancy status was ascertained from yes or no responses to the question “are you pregnant?”. Current lactation status was obtained from the question “are you still breastfeeding or giving breast milk to [your last child]”, which was posed to females aged 15-55 who reported having given birth in the preceding 5 years and having given their child breast milk. Participants who reported having given birth in the last five years answered additional questions about their use of folic acid-containing supplements in the three months prior to their most recent pregnancy, (“In the three months before you got pregnant with <baby’s name>, did you take a folic acid supplement or a multivitamin containing folic acid?”), and in the first three months of their most recent pregnancy (“During the first three months of your pregnancy with <baby’s name>, did you take a folic acid supplement or a multivitamin containing folic acid?), and about their awareness of the link between folic acid and birth defects (“Before your pregnancy with <baby’s name>, were you aware that taking folic acid before becoming pregnant can help prevent some birth defects?”).

*Other variables*

The categorization of other variables examined is as follows: highest level of education completed (≤ high school, postsecondary [trade, college, CEGEP or other non-university or university certificate or diploma below the bachelor's level], or university [bachelor's degree, diploma or degree above the bachelor's level]), household size-adjusted income (<median and ≥median, available for 2017/18 only, adjusted for household size using the square root of the number of persons in the household (2)), source of household income (employment [wages and salaries, income from self-employment, dividends and interest] and non-employment [employment insurance, worker's compensation, pensions, registered retirement savings plans or income funds, old-age security or guaranteed income supplement, social assistance or welfare, child support, alimony, other or none]), marital status (at time of survey; married [married or common-law], or other [single – never married, widowed, separated or divorced], immigration status (immigrated ≤5 years ago, immigrated >5 years ago, or born in Canada), racial group or Indigenous identity (self-reported, categorized according to Canadian Institute for Health Information standards (3): Black, East/Southeast Asian, First Nations, Inuit, Latin American, Métis, Middle Eastern, Multiple Indigenous, not stated, other or multiple races, South Asian, White), province or territory of residence (all Canadian provinces and territories), region of residence (Atlantic [New Brunswick, Newfoundland and Labrador, Nova Scotia, Prince Edward Island], Central [Ontario and Quebec], Northern [Yukon, Northwest Territories, Nunavut], Western [Alberta, British Columbia, Manitoba, Saskatchewan]), location of residence (urban or rural), food security (derived based on the food security module, and categorized as food secure, moderately food insecure, severely food insecure as described (4)), daily consumption of fruits and vegetables in the month prior to the survey (categorized as <5, ≥5 per day), monthly consumption of green vegetables in the month prior to the survey (categorized as <30, ≥30 per month), and frequency of general physical checkup (<every 3 years, ≥every 3 years or no pattern, available for 2015/16 only). CCHS annual included limited dietary intake data, but had

questions about daily consumption of fruits and vegetables in the month prior to the survey (categorized as <5, ≥5 per day), and monthly consumption of green vegetables in the month prior to the survey (categorized as <30, ≥30 per month). We included these questions in our analyses to capture the sources of folate that were available, and as markers of diet quality more broadly, as both are components of the Healthy Eating Index, and fruit and vegetable consumption specifically has been validated as an indicator of diet quality based on data from the 2004 CCHS - Nutrition (5). Respondent age at delivery was used for the peri-conception folic-acid containing supplement use, while respondent's current age was used for current supplement use (categorized ≤30 and >30 years). BMI (kg/m$^2$, categorized as underweight [<18.5], normal [18.5-24.9], overweight [25.0-29.9] and obese [≥30]), was calculated using self-reported pre-pregnancy weight and current self-reported height for the outcomes on folic acid-containing supplement use peri-conception among those who had given birth in the preceding 5 years, and using self-reported current height and weight for the question about current folic acid-containing supplement use among non-pregnant individuals. Pre-pregnancy BMI for the current pregnancy was not available for respondents who reported being pregnant at the time the survey was conducted. Respondents answered questions about frequency of alcohol use and smoking. Among those who had given birth in the last 5 years, for questions on retrospective folic acid supplement use in 3 months prior to and first 3 months of pregnancy, alcohol use and smoking were determined in the corresponding timeframes: in the 3 months prior to pregnancy, and after finding out about pregnancy, respectively. Alcohol use for each period was categorized as "yes" based on any frequency of alcohol consumption reported for the time frame examined, ("less than once per month", "once per month", "2 to 3 times per month", "once a week", 2 to 3 times per week", 4 to 6 times per week" or "everyday") and as "no" if the respondent reported no frequency of alcohol consumption for the period, or answered "no" to questions "Have you ever had a drink in your lifetime" and "During the past 12 months, that is from (date one year prior to interview), have you had a drink of beer, wine, liquor or any other alcoholic beverage?". Smoking was categorized as "yes" based on any frequency of smoking reported for the timeframe examined, ("everyday", "almost every day", "about 2 or 3 times a week", "about once a week", "Once or twice") or as "no" if the participant selected a frequency of "Never" or if the participant answered "not at all" to the question: "Do you smoke cigarettes daily, occasionally, or not at all?" and "no" to the question: "Have you smoked over 100 cigarettes in your lifetime?" For current folic acid supplement use for non-pregnant and pregnant women, respondents who answered yes to "drank alcohol - past week" were determined as current alcohol drinkers, and no if they answered no to this question or any drinks during lifetime and the past 12 months questions. They were noted as a current smoker if they answered they smoked cigarettes presently: "daily" or "occasionally", and a non-smoker if they answered "not at all".

#### Supplemental Results

**Supplemental Table 1.** Additional Characteristics - Folic acid-containing supplement use by among females aged 15-55 who are neither pregnant or lactating[1]

| **Subgroup** | **Sample Size[2]** | **Supplement Users** (% [95% CI]) | **Supplement Non-users** (% [95% CI ]) | **OR Unadjusted** ( 95% CI) | **OR Adjusted** (95% CI) |
|---|---|---|---|---|---|
| **Are you taking a vitamin supplement containing folic acid?" among those who report neither being currently pregnant or breastfeeding** | 53559 | 16.5 (15.9, 17.0) | 83.5 (83.0, 84.1) | n/a | n/a |
| **Source of Household income[3]** | | | | | |
| *Employment* | 22504 | 15.4 (14.7, 16.2) | 84.6 (83.8, 85.3) | n.s | n/a |
| Social assistance/welfare/alimony | 2220 | 16.3 (13.4, 19.3) | 83.7 (80.7, 86.6) | | |
| **Region of residence[4]** | | | | | |
| Atlantic | 6214 | 12.9 (11.7, 14.2) | 87.1 (85.8, 88.3) | **0.88 (0.78, 0.99)** | **1.14 (1.01, 1.29)** |
| *Central* | 26770 | 14.5 (13.8, 15.2) | 85.5 (84.8, 86.2) | Ref | Ref |
| Western | 19012 | 21.1 (20.3, 22.0) | 78.9 (78.0, 79.7) | **1.58 (1.46, 1.71)** | **2.20 (2.01, 2.42)** |
| Northern | 1563 | 14.9 (12.4, 17.4) | 85.1 (82.6, 87.6) | 1.04 (0.84, 1.27) | **1.64 (1.29, 2.09)** |
| **Type of community** | | | | | |
| Rural | 13322 | 14.5 (13.5, 15.5) | 85.5 (84.5, 86.5) | Ref | Ref |
| *Urban* | 40237 | 16.8 (16.2, 17.5) | 83.2 (82.5, 83.8) | **1.19 (1.09, 1.31)** | 1.09 (0.99, 1.20) |
| **Maternal pre-pregnancy BMI category** | | | | | |
| <18.5 | 2159 | 11.5 (9.4, 13.6) | 88.5 (86.4, 90.6) | **0.64 (0.52, 0.79)** | 0.80 (0.65, 0.99) [n.s global] |
| *18.5-24.9* | 25734 | 16.8 (16.1, 17.6) | 83.2 (82.4, 83.9) | Ref | Ref |

| | | | | | |
|---|---|---|---|---|---|
| 25-29.9 | 12722 | 16.6 (15.4, 17.7) | 83.4 (82.3, 84.6) | 0.98 (0.89, 1.08) | 0.91 (0.82, 1.00) |
| ≥30 | 10765 | 16.3 (15.1, 17.4) | 83.7 (82.6, 84.9) | 0.96 (0.87, 1.06) | 0.92 (0.82, 1.02) |
| **Food Security[5]** | | | | | |
| *Food secure* | 38031 | 15.7 (15.1, 16.2) | 84.3 (83.8, 84.9) | Ref | Ref |
| Moderately food insecure | 3877 | 14.6 (12.7, 16.4) | 85.4 (83.6, 87.3) | 0.92 (0.79, 1.07) | 1.11 (0.94, 1.31) |
| Severely food insecure | 2194 | 14.4 (12.1, 16.7) | 85.6 (83.3, 87.9) | 0.91 (0.74, 1.11) | 1.24 (1.00, 1.53) |
| **Consumption of green vegetables in the month prior to survey[6]** | | | | | |
| less than 30 per month | 13141 | 15.0 (13.9, 16.1) | 85.0 (83.9, 86.1) | **0.73 (0.66, 0.81)** | 0.88 (0.78, 0.99) [n.s global] |
| *30 or more per month* | 14186 | 19.5 (18.5, 20.6) | 80.5 (79.4, 81.5) | Ref | Ref |
| **Daily consumption fruits and vegetables in month prior to survey[6]** | | | | | |
| less than 5 | 17007 | 15.7 (14.7, 16.7) | 84.3 (83.3, 85.3) | **0.74 (0.66, 0.82)** | **0.78 (0.69, 0.88)** |
| *5 or more* | 9896 | 20.2 (18.9, 21.5) | 79.8 (78.5, 81.1) | Ref | Ref |
| **Frequency of general physical check-up[7]** | | | | | |
| *Less than every 3 years* | 4281 | 8.6 (7.3, 10.0) | 91.4 (90.0, 92.7) | Ref | Ref |
| Every 3 year or more or no regulat pattern | 1347 | 5.5 (3.9, 7.1) | 94.5 (92.9, 96.1) | **0.62 (0.44, 0.87)** | n/a |
| **Alcohol use in the past week** | | | | | |
| Drank | 18406 | 17.3 (16.4, 18.2) | 82.7 (81.8, 83.6) | n.s | n/a |
| *Did not drink* | 19968 | 17.8 (16.8, 18.7) | 82.2 (81.3, 83.2) | | |
| **Current smoking status** | | | | | |
| Daily | 8149 | 11.4 (10.4, 12.5) | 88.6 (87.5, 89.6) | **0.62 (0.56, 0.70)** | **0.73 (0.65, 0.83)** |
| Occasionally | 2881 | 16.4 (14.2, 18.6) | 83.6 (81.4, 85.8) | 0.95 (0.80, 1.12) | 1.14 (0.96, 1.35) |
| *Not at all* | 42517 | 17.2 (16.6, 17.8) | 82.8 (82.2, 83.4) | Ref | Ref |

1 All characteristics refer to current status at the time the survey was conducted, except where noted otherwise. Multivariable models included all factors significantly associated with folic acid-containing supplement use in the univariate model; factors whose global association was not significant were excluded and denoted with n.s. Separate models were run for income and education, and for province/territory of residence and region of residence to avoid collinearity. Bold indicates significant differences (p<0.05) from the reference group after adjustment for multiple comparisons. Superscript E indicates that values should be interpreted with caution due to a wide CV (16.6-33.3%), while estimates replaced by an F are suppressed due to low sample size (<10) and/or a CV ≥33.3%.

2 Sample sizes refer to those who answered either yes or no. Sample sizes and proportions may not add to 100%/the full sample size because some responses were do not know or refuse to answer, or because question was not included for all cycles/and/or provinces/territories as specified below.

3 Total household income before taxes adjusted for household size (income/square root of size)(2). Income available in this format only for 2017/18 cycle.

4 Categorized as Atlantic [New Brunswick, Newfoundland and Labrador, Nova Scotia, Prince Edward Island], Central [Ontario and Quebec], Northern [Yukon, Northwest Territories, Nunavut], Western [Alberta, British Columbia, Manitoba, Saskatchewan].

5 Module was optional content for 2015-16 for NFLD and Labrador, Ontario, module not included for Yukon. The global p-value was significant in the unadjusted model though the pairwise comparisons shown were not, so it was also examined in the adjusted model.

6 Module was core content for 2015/16 only.

7 Question included as optional content in the 2015/16 survey only. Excluded from multivariable model as it was asked in less than 50% of respondents.

**Supplemental Table 2.** Additional Characteristics - Folic acid-containing supplement use by lactating females aged 15-55[1]

| **Subgroup** | **Sample Size**[2] | **Supplement Users** (% [95% CI]) | **Supplement Non-users** (% [95% CI]) | **OR Unadjusted** (95% CI) | **OR Adjusted** (95% CI) |
|---|---|---|---|---|---|
| **Are you taking a vitamin supplement containing folic acid?" among those who report not being pregnant and currently breastfeeding** | 2069 | 58.4 (54.8, 61.9) | 41.6 (38.1, 45.2) | n/a | n/a |
| **Source of Household income**[3] | | | | | |
| *Employment* | 909 | 59.0 (53.6, 64.3) | 41.0 (35.7, 46.4) | n.s | n/a |
| Social assistance/welfare/alimony | 69 | 41.2 (19.4, 63.0) E | 58.8 (37.0, 80.6) E | | |
| **Region of residence**[4] | | | | | |
| Atlantic | 151 | 59.9 (49.5, 70.3) | 40.1 (29.7, 50.5) | 1.12 (0.69, 1.79) | 1.64 (0.97, 2.78) |
| *Central* | 899 | 57.3 (52.2, 62.4) | 42.7 (37.6, 47.8) | Ref | Ref |
| Western | 892 | 60.4 (55.8, 65) | 39.6 (35.0, 44.2) | 1.14 (0.86, 1.51) | **1.56 (1.15, 2.13)** |
| Northern | 127 | 34.4 (21.9, 47.0) E | 65.6 (53.0, 78.1) | **0.39 (0.22, 0.71)** | 0.91 (0.46, 1.79) |
| **Type of community** | | | | | |
| Rural | 472 | 53.6 (47.0, 60.3) | 46.4 (39.7, 53.0) | n.s | n/a |
| *Urban* | 1597 | 59.1 (55.2, 63.1) | 40.9 (36.9, 44.8) | | |
| **Maternal pre-pregnancy BMI category** | | | | | |
| <18.5 | 83 | 57.3 (42.0, 72.5) | 42.7 (27.5, 58.0) E | n.s | n/a |
| *18.5-24.9* | 1066 | 59.5 (54.7, 64.3) | 40.5 (35.7, 45.3) | | |
| 25-29.9 | 542 | 58.4 (51.8, 65.1) | 41.6 (34.9, 48.2) | | |

| | | | | | |
|---|---|---|---|---|---|
| ≥30 | 314 | 54.4 (44.4, 64.4) | 45.6 (35.6, 55.6) | | |
| **Food Security[5]** | | | | | |
| *Food secure* | 1532 | 58.7 (54.8, 62.6) | 41.3 (37.4, 45.2) | Ref | Ref |
| Moderately food insecure | 157 | 42.7 (28.6, 56.7) E | 57.3 (43.3, 71.4) | **0.52 (0.29, 0.96)** | 0.53 (0.26, 1.07) |
| Severely food insecure | 48 | F | 69.0 (44.1, 94.0) E | 0.32 (0.09, 1.08) | 0.49 (0.13, 1.77) |
| **Consumption of green vegetables in the month prior to survey[6]** | | | | | |
| less than 30 per month | 445 | 59.0 (51.5, 66.5) | 41.0 (33.5, 48.5) | n.s | n/a |
| *30 or more per month* | 654 | 58.7 (52.3, 65.1) | 41.3 (34.9, 47.7) | | |
| **Daily consumption fruits and vegetables in month prior to survey[6]** | | | | | |
| less than 5 | 568 | 64.1 (57.9, 70.4) | 35.9 (29.6, 42.1) | **1.57 (1.04, 2.36)** | **1.57 (1.01, 2.45)** |
| *5 or more* | 512 | 53.3 (45.8, 60.7) | 46.7 (39.3, 54.2) | Ref | Ref |
| **Frequency of general physical check-up[7]** | | | | | |
| *Less than every 3 years* | 108 | 41.7 (28.4, 55.1) | 58.3 (44.9, 71.6) | n.s | n/a |
| Every 3 year or more or no regulat pattern | 45 | 44.2 (25.4, 63.0) E | 55.8 (37.0, 74.6) E | | |
| **Alcohol use in the past week** | | | | | |
| Drank | 527 | 52.4 (45.9, 58.8) | 47.6 (41.2, 54.1) | **0.60 (0.42, 0.84)** | **0.59 (0.41, 0.85)** |
| *Did not drink* | 958 | 64.9 (59.6, 70.2) | 35.1 (29.8, 40.4) | Ref | Ref |
| **Current smoking status** | | | | | |
| Daily | 150 | 39.7 (24.8, 54.5) E | 60.3 (45.5, 75.2) | **0.44 (0.23, 0.87)** | 0.64 (0.33, 1.23) |
| Occasionally | 72 | 41.3 (22.7, 59.9) E | 58.7 (40.1, 77.3) | 0.48 (0.21, 1.06) | 0.66 (0.30, 1.44) |
| *Not at all* | 1845 | 59.7 (56.0, 63.4) | 40.3 (36.6, 44.0) | Ref | Ref |

1 All characteristics refer to current status at the time the survey was conducted, except where noted otherwise. Multivariable models included all factors significantly associated with folic acid-containing supplement use in the univariate model; factors whose global association was not significant were excluded and denoted with n.s. Separate models were run for income and education, and for province/territory of residence and region of residence to avoid collinearity. Bold indicates significant differences ($p<0.05$) from the reference group after adjustment for multiple comparisons. Superscript E indicates that values should be interpreted with caution due to a wide CV (16.6-33.3%), while estimates replaced by an F are suppressed due to low sample size (<10) and/or a CV ≥33.3%.

2 Sample sizes refer to those who answered either yes or no. Sample sizes and proportions may not add to 100%/the full sample size because some responses were do not know or refuse to answer, or because question was not included for all cycles/and/or provinces/territories as specified below.

3 Total household income before taxes adjusted for household size (income/square root of size)(2). Income available in this format only for 2017/18 cycle.

4 Categorized as Atlantic [New Brunswick, Newfoundland and Labrador, Nova Scotia, Prince Edward Island], Central [Ontario and Quebec], Northern [Yukon, Northwest Territories, Nunavut], Western [Alberta, British Columbia, Manitoba, Saskatchewan].

5 Module was optional content for 2015-16 for NFLD and Labrador, Ontario, module not included for Yukon.

6 Module was core content for 2015/16 only.

7 Question included as optional content in the 2015/16 survey only.

**Supplemental Table 3.** Additional Characteristics - Folic acid-containing supplement use among pregnant females aged 15-55[1].

| Subgroup | Sample Size[2] | Supplement Users (% [95% CI]) | Supplement Non-users (% [95% CI]) | OR Unadjusted (95% CI) | OR Adjusted (95% CI) |
|---|---|---|---|---|---|
| **"Are you taking a vitamin supplement containing folic acid?" among those who report being currently pregnant** | 1676 | 80.3 (77.1, 83.5) | 19.7 (16.5, 22.9) | n/a | n/a |
| **Source of Household income[3]** | | | | | |
| *Employment* | 711 | 83.1 (78.8, 87.5) | 16.9 (12.5, 21.2) | Ref | n/a |
| Social assistance/welfare/alimony | 63 | 71.8 (56.8, 86.8) | 28.2 (13.2, 43.2) E | 0.52 (0.23, 1.14) | |
| **Region of residence[4]** | | | | | |
| Atlantic | 154 | 90.3 (85.8, 94.8) | 9.7 (5.2, 14.2) E | **2.82 (1.55, 5.12)** | 2.58 (1.26, 5.26) [n.s global] |
| *Central* | 829 | 76.8 (72.1, 81.5) | 23.2 (18.5, 27.9) | Ref | Ref |
| Western | 631 | 85.2 (81.5, 89.0) | 14.8 (11.0, 18.5) | **1.75 (1.18, 2.58)** | 1.35 (0.74, 2.47) |
| Northern | 62 | 77.1 (64.1, 90.2) | 22.9 (9.8, 35.9) E | 1.02 (0.45, 2.29) | 3.26 (0.49, 21.90) |
| **Type of community** | | | | | |
| Rural | 423 | 79.6 (74.2, 85.0) | 20.4 (15.0, 25.8) | n.s | n/a |
| *Urban* | 1253 | 80.5 (76.8, 84.1) | 19.5 (15.9, 23.2) | | |
| **Food Security[5]** | | | | | |
| *Food secure* | 1180 | 83.4 (79.7, 87.0) | 16.6 (13.0, 20.3) | Ref | Ref |
| Moderately food insecure | 141 | 70.1 (59.2, 81.1) | 29.9 (18.9, 40.8) E | **0.47 (0.26, 0.86)** | 0.75 (0.35, 1.57) |
| Severely food insecure | 61 | 74.9 (58.6, 91.2) | 25.1 (8.8, 41.4) E | 0.60 (0.22, 1.65) | 0.90 (0.16, 4.96) |

| | | | | | |
|---|---|---|---|---|---|
| **Consumption of green vegetables[6]** | | | | | |
| <median (<30 per month) | 429 | 80.1 (74.0, 86.1) | 19.9 (13.9, 26.0) | n.s | n/a |
| *≥median (≥30 per month)* | 459 | 81.0 (75.4, 86.6) | 19.0 (13.4, 24.6) | | |
| **Consumption of total fruits and vegetables[6]** | | | | | |
| less than median (<5 per day) | 446 | 73.0 (66.2, 79.8) | 27.0 (20.2, 33.8) | **0.40 (0.24, 0.68)** | 0.49 (0.25, 0.95) [n.s global] |
| *More than median (≥5 per day)* | 432 | 87.1 (82.3, 92.0) | 12.9 (8.0, 17.7) E | Ref | Ref |
| **Frequency of general physical check-up[7]** | | | | | |
| *Less than every 3 years* | 140 | 81.0 (71.4, 90.6) | 19.0 (9.4, 28.6) E | n.s | n/a |
| Every 3 year or more or no regular pattern | 43 | 83.8 (72.3, 95.2) | F | | |
| **Alcohol use in the past week** | | | | | |
| Drank | 94 | 23.1 (11.8, 34.4) E | 76.9 (65.6, 88.2) | **0.06 (0.03, 0.12)** | **0.05 (0.02, 0.11)** |
| *Did not drink* | 1103 | 83.1 (79.3, 86.9) | 16.9 (13.1, 20.7) | Ref | Ref |
| **Smoking** | | | | | |
| Daily | 143 | 58.5 (44.3, 72.6) | 41.5 (27.4, 55.7) E | **0.31 (0.16, 0.59)** | 0.74 (0.31, 1.76) |
| Occasionally | 60 | 68.1 (48.3, 88.0) | 31.9 (12.0, 51.7) E | 0.47 (0.18, 1.26) | 0.75 (0.24, 2.35) |
| *Not at all* | 1473 | 82.0 (78.7, 85.2) | 18.0 (14.8, 21.3) | Ref | Ref |

1 All characteristics refer to current status at the time the survey was conducted, except where noted otherwise. Multivariable models included all factors significantly associated with folic acid-containing supplement use in the univariate model; factors whose global association was not significant were excluded and denoted with n.s. Separate models were run for income and education, and for province/territory of residence and region of residence to avoid collinearity. Bold indicates significant differences (p<0.05) from the reference group after adjustment for multiple comparisons. Superscript E indicates that values should be interpreted with caution due to a wide CV (16.6-33.3%), while estimates replaced by an F are suppressed due to low sample size (<10) and/or a CV ≥33.3%. Pre-pregnancy BMI was unavailable for currently pregnant respondents, and was therefore not included in the table.

2 Sample sizes refer to those who answered either yes or no. Sample sizes and proportions may not add to 100%/the full sample size because some responses were do not know or refuse to answer, or because question was not included for all cycles/and/or provinces/territories as specified below.

3 Total household income before taxes adjusted for household size (income/square root of size)(2). Income available in this format only for 2017/18 cycle.

4 Categorized as Atlantic [New Brunswick, Newfoundland and Labrador, Nova Scotia, Prince Edward Island], Central [Ontario and Quebec], Northern [Yukon, Northwest Territories, Nunavut], Western [Alberta, British Columbia, Manitoba, Saskatchewan].

5 Module was optional content for 2015-16 for NFLD and Labrador, Ontario, module not included for Yukon.

6 Module was core content for 2015/16 only.

7 Question included as optional content in the 2015/16 survey only.

**Supplemental Table 4.** Additional Characteristics - Awareness of link between folic acid and some birth defects[1].

| **Subgroup** | **Sample Size[2]** | **Aware** (% [95% CI]) | **Not-Aware** (% [95% CI ]) | **OR Unadjusted** (95% CI) | **OR Adjusted** (95% CI) |
|---|---|---|---|---|---|
| "Before your pregnancy with <baby's name>, were you aware that taking folic acid before becoming pregnant can help prevent some birth defects?" | 10384 | 76.3 (74.9, 77.6) | 23.7 (22.4, 25.1) | n/a | n/a |
| **Source of Household income[3]** | | | | | |
| Employment | 4476 | 81.1 (79.3, 82.8) | 18.9 (17.2, 20.7) | Ref. | Ref. |
| Social assistance/welfare/alimony/other | 531 | 50.2 (41.8, 58.6) | 49.8 (41.4, 58.2) | **0.24 (0.17, 0.34)** | **0.53 (0.34, 0.82)** |
| **Region of residence[4]** | | | | | |
| Atlantic | 983 | 75.9 (72.3, 79.5) | 24.1 (20.5, 27.7) | 0.90 (0.72, 1.13) | 0.85 (0.66, 1.09) |
| *Central* | 5028 | 77.8 (75.9, 79.6) | 22.2 (20.4, 24.1) | Ref. | Ref. |
| Western | 3957 | 73.9 (71.9, 76.0) | 26.1 (24.0, 28.1) | **0.81 (0.70, 0.95)** | 0.83 (0.69, 1.01) |
| Northern | 416 | 52.6 (47.1, 58.0) | 47.4 (42.0, 52.9) | **0.32 (0.25, 0.41)** | 0.61 (0.35, 1.06) |
| **Type of community** | | | | | |
| *Rural* | 2725 | 79.7 (77.6, 81.9) | 20.3 (18.1, 22.4) | Ref | Ref |
| Urban | 7659 | 75.6 (74.0, 77.1) | 24.4 (22.9, 26.0) | **0.79 (0.67, 0.92)** | 0.98 (0.81, 1.19) |
| **Maternal pre-pregnancy BMI category** | | | | | |
| <18.5 | 491 | 69.0 (63.0, 75.0) | 31.0 (25.0, 37.0) | **0.61 (0.45, 0.83)** | 0.83 (0.60, 1.15) |
| *18.5-24.9* | 5281 | 78.5 (76.6, 80.4) | 21.5 (19.6, 23.4) | Ref. | Ref. |
| 25-29.9 | 2062 | 77.0 (74.0, 80.0) | 23.0 (20.0, 26.0) | 0.92 (0.75, 1.12) | 0.93 (0.75, 1.16) |
| ≥30 | 1332 | 75.4 (71.3, 79.5) | 24.6 (20.5, 28.7) | 0.84 (0.66, 1.07) | 0.94 (0.71, 1.24) |
| **Food Security[5]** | | | | | |
| *Food secure* | 7340 | 79.8 (78.4, 81.3) | 20.2 (18.7, 21.6) | Ref. | Ref. |
| Moderately food insecure | 962 | 58.6 (53.1, 64.1) | 41.4 (35.9, 46.9) | **0.36 (0.28, 0.46)** | **0.66 (0.49, 0.87)** |
| Severely food insecure | 414 | 56.8 (48.7, 65.0) | 43.2 (35.0, 51.3) | **0.33 (0.24, 0.47)** | **0.61 (0.41, 0.90)** |
| **Consumption of total fruits and vegetables[6]** | | | | | |

| | | | | | |
|---|---|---|---|---|---|
| <5 per day | 3017 | 70.7 (67.8, 73.5) | 29.3 (26.5, 32.2) | **0.59 (0.48, 0.73)** | **0.76 (0.57, 1.00)** |
| >5 per day | 2230 | 80.4 (77.9, 82.9) | 19.6 (17.1, 22.1) | Ref. | Ref. |
| **Consumption of green vegetables[6]** | | | | | |
| <30 per month | 2385 | 71.8 (68.7, 74.9) | 28.2 (25.1, 31.3) | **0.76 (0.62, 0.94)** | 1.00 (0.78, 1.29) |
| *>30 per month* | 2932 | 76.9 (74.5, 79.4) | 23.1 (20.6, 25.5) | Ref. | Ref. |
| **Frequency of general physical check-up[7]** | | | | | |
| *Less than every 3 years* | 755 | 82.0 (78.0, 86.0) | 18.0 (14.0, 22.0) | n.s | n/a |
| Every 3 year or more or no regular pattern | 247 | 76.9 (68.7, 85.1) | 23.1 (14.9, 31.3) E | | |
| **Alcohol use in 3 months prior to pregnancy[6]** | | | | | |
| Drank | 2608 | 81.7 (79.7, 83.8) | 18.3 (16.2, 20.3) | **1.54 (1.26, 1.88)** | 1.02 (0.80, 1.31) |
| *Did not drink* | 2585 | 74.4 (71.6, 77.2) | 25.6 (22.8, 28.4) | Ref. | Ref. |
| **Smoking in the 3 months prior to pregnancy[6]** | | | | | |
| Smoked | 1030 | 61.4 (56.4, 66.3) | 38.6 (33.7, 43.6) | **0.38 (0.30, 0.48)** | **0.48 (0.35, 0.64)** |
| *Did not smoke* | 4201 | 80.7 (78.8, 82.6) | 19.3 (17.4, 21.2) | Ref. | Ref. |
| **Years since birth of most recent infant** | | | | | |
| 0 | 2417 | 79.8 (77.2, 82.4) | 20.2 (17.6, 22.8) | Ref. | |
| 1 | 2370 | 77.8 (75.2, 80.5) | 22.2 (19.5, 24.8) | 0.89 (0.71, 1.12) | 1.02 (0.78, 1.33) |
| 2 | 1926 | 79.3 (76.4, 82.1) | 20.7 (17.9, 23.6) | 0.97 (0.77, 1.22) | 1.17 (0.90, 1.51) |
| 3 | 1674 | 73.0 (69.4, 76.6) | 27.0 (23.4, 30.6) | **0.68 (0.54, 0.88)** | 0.81 (0.61, 1.08) |
| 4 | 1529 | 69.6 (65.7, 73.5) | 30.4 (26.5, 34.3) | **0.58 (0.46, 0.74)** | **0.66 (0.50, 0.88)** |
| 5 | 468 | 69.0 (61.0, 76.9) | 31.0 (23.1, 39.0) | **0.56 (0.37, 0.85)** | 0.73 (0.47, 1.15) |

1 All characteristics refer to current status at the time the survey was conducted, except where noted otherwise. Multivariable models included all factors significantly associated with folic acid-containing supplement use in the univariate model; factors whose global association was not significant were excluded and denoted with n.s. Separate models were run for income and education, and for province/territory of residence and region of residence to avoid collinearity. Bold indicates significant differences (p<0.05) from the reference group after adjustment for multiple comparisons. Superscript E indicates that values should be interpreted with caution due to a wide CV (16.6-33.3%), while estimates replaced by an F are suppressed due to low sample size (<10) and/or a CV >33.3%.

2 Sample sizes refer to those who answered either yes or no. Sample sizes and proportions may not add to 100%/the full sample size because some responses were do not know or refuse to answer, or because question was not included for all cycles/and/or provinces/territories as specified below.

3 Total household income before taxes adjusted for household size (income/square root of size)(2). Income available in this format only for 2017/18 cycle.

4 Categorized as Atlantic [New Brunswick, Newfoundland and Labrador, Nova Scotia, Prince Edward Island], Central [Ontario and Quebec], Northern [Yukon, Northwest Territories, Nunavut], Western [Alberta, British Columbia, Manitoba, Saskatchewan].

5 Module was optional content for 2015-16 for NFLD and Labrador, Ontario, module not included for Yukon.

6 Module was core content for 2015/16 only.

7 Question included as optional content in the 2015/16 survey only.

**Supplemental Table 5.** Additional Characteristics - Folic acid-containing supplement in the 3 months prior to becoming pregnant[1].

| **Subgroup** | **Sample Size**[2] | **Supplement Users** (% [95% CI]) | **Supplement Non-users** (% [95% CI ]) | **OR Unadjusted** (95% CI) | **OR Adjusted** (95% CI) |
|---|---|---|---|---|---|
| **“In the three months before you got pregnant with <baby’s name>, did you take a folic acid supplement or a multivitamin containing folic acid?”** | 10326 | 63.7 (62.2-65.1) | 36.3 (34.9-37.8) | n/a | N/a |
| **Source of Household income**[3] | | | | | |
| Employment | 4461 | 66.2 (64.1, 68.4) | 33.8 (31.6, 35.9) | Ref | Ref |
| Social assistance/welfare/alimony/other | 531 | 39.4 (31.5, 47.3) | 60.6 (52.7, 68.5) | **0.33 (0.24, 0.47)** | 0.80 (0.52, 1.24) |
| **Region of residence**[4] | | | | | |
| Atlantic | 981 | 55.0 (50.6, 59.4) | 45.0 (40.6, 49.4) | **0.63 (0.52, 0.77)** | **0.67 (0.53, 0.84)** |
| *Central* | 5000 | 65.8 (63.8, 67.9) | 34.2 (32.1, 36.2) | Ref | Ref |
| Western | 3935 | 61.6 (59.4, 63.8) | 38.4 (36.2, 40.6) | **0.83 (0.73, 0.95)** | 0.90 (0.71, 1.14) |
| Northern | 410 | 42.1 (35.8, 48.3) | 57.9 (51.7, 64.2) | **0.38 (0.29, 0.49)** | 0.87 (0.74, 1.04) |
| **Type of community** | | | | | |
| *Rural* | 2708 | 63.6 (60.9, 66.3) | 36.4 (33.7, 39.1) | n.s | n/a |
| Urban | 7618 | 63.7 (62.0, 65.4) | 36.3 (34.6, 38.0) | | |
| **Maternal pre-pregnancy BMI category** | | | | | |
| <18.5 | 487 | 55.7 (48.9, 62.5) | 44.3 (37.5, 51.1) | **0.61 (0.45, 0.81)** | **0.71 (0.51, 0.98)** |
| *18.5-24.9* | 5254 | 67.5 (65.5, 69.4) | 32.5 (30.6, 34.5) | Ref | Ref |
| 25-29.9 | 2049 | 62.5 (59.1, 65.9) | 37.5 (34.1, 40.9) | **0.80 (0.68, 0.96)** | 0.84 (0.69, 1.02) |
| ≥30 | 1328 | 55.7 (51.2, 60.2) | 44.3 (39.8, 48.8) | **0.61 (0.49, 0.75)** | **0.71 (0.56, 0.89)** |
| **Food Security**[5] | | | | | |

| | | | | | |
|---|---|---|---|---|---|
| *Food secure* | 7298 | 66.9 (65.3, 68.6) | 33.1 (31.4, 34.7) | Ref | Ref |
| Moderately food insecure | 955 | 45.9 (40.2, 51.7) | 54.1 (48.3, 59.8) | **0.42 (0.33, 0.54)** | 0.79 (0.60, 1.04) |
| Severely food insecure | 413 | 39.9 (31.8, 47.9) | 60.1 (52.1, 68.2) | **0.33 (0.23, 0.47)** | 0.71 (0.45, 1.11) |
| **Consumption of green vegetables**[6] | | | | | |
| <30 per month | 2370 | 59.4 (56.2, 62.6) | 40.6 (37.4, 43.8) | **0.72 (0.59, 0.86)** | 0.84 (0.67, 1.05) |
| *≥30 per month* | 2906 | 67.2 (64.4, 69.9) | 32.8 (30.1, 35.6) | Ref | Ref |
| **Consumption of total fruits and vegetables**[6] | | | | | |
| <5 per day | 3002 | 60.1 (57.2, 62.9) | 39.9 (37.1, 42.8) | **0.66 (0.54, 0.80)** | 0.93 (0.72, 1.18) |
| >5 per day | 2206 | 69.6 (66.5, 72.7) | 30.4 (27.3, 33.5) | Ref | Ref |
| **Frequency of general physical check-up**[7] | | | | | |
| *Less than every 3 years* | 751 | 70.5 (65.9, 75.1) | 29.5 (24.9, 34.1) | n.s | n/a |
| Every 3 year or more or no regular pattern | 246 | 63.6 (54.5, 72.8) | 36.4 (27.2, 45.5) | | |
| **Alcohol use in 3 months prior to pregnancy**[6] | | | | | |
| Drank | 2602 | 62.6 (59.8, 65.5) | 37.4 (34.5, 40.2) | n.s | n/a |
| *Did not drink* | 2571 | 64.2 (61.1, 67.4) | 35.8 (32.6, 38.9) | Ref | Ref |
| **Smoking in the 3 months prior to pregnancy**[6] | | | | | |
| Smoked | 1024 | 43.0 (38.0, 47.9) | 57.0 (52.1, 62.0) | **0.37 (0.29, 0.46)** | **0.62 (0.46, 0.84)** |
| *Did not smoke* | 4188 | 67.2 (64.9, 69.5) | 32.8 (30.5, 35.1) | Ref | Ref |
| **Years since birth of most recent infant** | | | | | |
| 0 | 2410 | 66.7 (63.7, 69.7) | 33.3 (30.3, 36.3) | n.s | n/a |
| 1 | 2367 | 63.7 (60.6, 66.8) | 36.3 (33.2, 39.4) | | |
| 2 | 1909 | 64.0 (60.5, 67.5) | 36.0 (32.5, 39.5) | | |

| | | | |
|---|---|---|---|
| 3 | 1662 | 60.5 (56.8, 64.3) | 39.5 (35.7, 43.2) |
| 4 | 1514 | 62.8 (59.3, 66.3) | 37.2 (33.7, 40.7) |
| 5 | 464 | 59.4 (51.0, 67.9) | 40.6 (32.1, 49.0) |

1 All characteristics refer to current status at the time the survey was conducted, except where noted otherwise. Multivariable models included all factors significantly associated with folic acid-containing supplement use in the univariate model; factors whose global association was not significant were excluded and denoted with n.s. Separate models were run for income and education, and for province/territory of residence and region of residence to avoid collinearity. Bold indicates significant differences ($p<0.05$) from the reference group after adjustment for multiple comparisons. Superscript E indicates that values should be interpreted with caution due to a wide CV (16.6-33.3%), while estimates replaced by an F are suppressed due to low sample size (<10) and/or a CV ≥33.3%.

2 Sample sizes refer to those who answered either yes or no. Proportions may not add to 100%/the full sample size because some responses were do not know or refuse to answer, or because question was not included for all cycles/and/or provinces/territories as specified below.

3 Total household income before taxes adjusted for household size (income/square root of size)(2). Income available in this format only for 2017/18 cycle.

4 Categorized as Atlantic [New Brunswick, Newfoundland and Labrador, Nova Scotia, Prince Edward Island], Central [Ontario and Quebec], Northern [Yukon, Northwest Territories, Nunavut], Western [Alberta, British Columbia, Manitoba, Saskatchewan].

5 Module was optional content for 2015-16 for NFLD and Labrador, Ontario, module not included for Yukon.

6 Module was core content for 2015/16 only.

7 Question included as optional content in the 2015/16 survey only.

**Supplemental Table 6.** Additional Characteristics - Folic acid-containing supplement use in the first three months of pregnancy[1].

| **Subgroup** | **Sample Size[2]** | **Supplement Users (% [95% CI])** | **Supplement Non-users (% [95% CI ])** | **OR Unadjusted** (95% CI) | **OR Adjusted** (95% CI) |
|---|---|---|---|---|---|
| **"During the first three months of your pregnancy with <baby's name, did you take a folic acid supplement or a multivitamin containing folic acid?"** | 10329 | 89.9 (88.8, 90.9) | 10.1 (9.1, 11.2) | | |
| **Source of Household income[3]** | | | | | |
| *Employment* | 4459 | 91.1 (89.7, 92.6) | 8.9 (7.4, 10.3) | Ref | Ref |
| Social assistance/welfare/alimony/other | 533 | 72.3 (63.6, 81.0) | 27.7 (19.0, 36.4) | **0.25 (0.16, 0.41)** | 0.62 (0.35, 1.12) |
| **Region of residence[4]** | | | | | |
| Atlantic | 982 | 90.8 (88.4, 93.3) | 9.2 (6.7, 11.6) | 1.19 (0.85, 1.66) | 1.38 (0.95, 2.00) |
| *Central* | 5001 | 89.3 (87.8, 90.8) | 10.7 (9.2, 12.2) | Ref | Ref |
| Western | 3932 | 90.9 (89.6, 92.2) | 9.1 (7.8, 10.4) | 1.20 (0.95, 1.50) | **1.45 (1.13, 1.87)** |
| Northern | 414 | 76.6 (71.0, 82.2) | 23.4 (17.8, 29.0) | **0.39 (0.28, 0.56)** | 1.26 (0.63, 2.54) |
| **Type of community** | | | | | |
| *Rural* | 2714 | 90.6 (89.0, 92.2) | 9.4 (7.8, 11.0) | n.s | n/a |
| *Urban* | 7615 | 89.7 (88.5, 90.9) | 10.3 (9.1, 11.5) | | |
| **Maternal pre-pregnancy BMI category** | | | | | |
| <18.5 | 485 | 87.5 (83.4, 91.7) | 12.5 (8.3, 16.6) E | 0.72 (0.47, 1.09) | 0.98 (0.62, 1.55) |
| *18.5-24.9* | 5253 | 90.7 (89.3, 92.1) | 9.3 (7.9, 10.7) | Ref | Ref |
| 25-29.9 | 2052 | 91.3 (89.4, 93.1) | 8.7 (6.9, 10.6) | 1.07 (0.80, 1.43) | 1.13 (0.84, 1.51) |
| $\geq$30 | 1326 | 85.2 (81.0, 89.3) | 14.8 (10.7, 19) | **0.59 (0.41, 0.85)** | **0.68 (0.46, <1.00)** |

| | | | | | |
|---|---|---|---|---|---|
| **Food Security[5]** | | | | | |
| *Food secure* | 7297 | 90.8 (89.7, 92.0) | 9.2 (8.0, 10.3) | Ref | Ref |
| Moderately food insecure | 958 | 79.8 (75.4, 84.2) | 20.2 (15.8, 24.6) | **0.40 (0.29, 0.55)** | 0.75 (0.53, 1.07) |
| Severely food insecure | 411 | 81.4 (74.9, 87.9) | 18.6 (12.1, 25.1) E | **0.44 (0.28, 0.71)** | 0.86 (0.52, 1.43) |
| **Consumption of green vegetables[6]** | | | | | |
| <30 per month | 2367 | 86.6 (84.1, 89.2) | 13.4 (10.8-15.9) | **0.48 (0.36, 0.64)** | **0.53 (0.36, 0.77)** |
| *≥30 per month* | 2913 | 93.1 (92.0, 94.3) | 6.7 (5.7-8.0) | Ref | Ref |
| **Consumption of total fruits and vegetables[6]** | | | | | |
| <5 per day | 2996 | 88.7 (86.7, 90.7) | 11.3 (9.3, 13.3) | **0.64 (0.47, 0.88)** | 1.21 (0.81, 1.81) |
| *≥5 per day* | 2216 | 92.5 (90.8, 94.2) | 7.5 (5.8, 9.2) | Ref | Ref |
| **Frequency of general physical check-up[7]** | | | | | |
| *Less than every 3 years* | 753 | 88.9 (85.3, 92.5) | 11.1 (7.5, 14.7) E | n.s | n/a |
| Every 3 year or more or no regular pattern | 243 | 83.3 (74.8, 91.9) | 16.7 (8.1, 25.2) E | | |
| **Alcohol use in the first 3 months of pregnancy[6]** | | | | | |
| Drank | 213 | 91.1 (86.8, 95.3) | 8.9 (4.7, 13.2) E | n.s | n/a |
| *Did not drink* | 5003 | 89.3 (87.8, 90.9) | 10.7 (9.1, 12.2) | | |
| **Smoking in the first 3 months of pregnancy[6]** | | | | | |
| Smoked | 604 | 78.1 (72.0, 84.2) | 21.9 (15.8, 28) | **0.38 (0.25, 0.57)** | **0.62 (0.39, 0.98)** |
| *Did not smoke* | 4606 | 90.5 (88.9, 92.0) | 9.5 (8.0, 11.1) | Ref | Ref |
| **Years since birth of most recent infant** | | | | | |
| *0* | 2410 | 92.4 (90.6, 94.1) | 7.6 (5.9, 9.4) | | |
| 1 | 2362 | 89.5 (87.3, 91.7) | 10.5 (8.3, 12.7) | 0.71 (0.50, 0.99) | 0.73 (0.52, 1.05) |

| | | | | | |
|---|---|---|---|---|---|
| 2 | 1912 | 89.2 (86.8, 91.6) | 10.8 (8.4, 13.2) | 0.68 (0.47, 0.98) | 0.76 (0.52, 1.10) |
| 3 | 1664 | 88.6 (85.8, 91.5) | 11.4 (8.5, 14.2) | **0.65 (0.44, 0.95)** | 0.76 (0.53, 1.10) |
| 4 | 1521 | 89.9 (87.9, 92.0) | 10.1 (8.0, 12.1) | 0.74 (0.53, 1.04) | 1.01 (0.69, 1.47) |
| 5 | 460 | 84.9 (78.0, 91.9) | 15.1 (8.1, 22) E | 0.47 (0.25, 0.85) | 0.69 (0.41, 1.17) |

1 All characteristics refer to current status at the time the survey was conducted, except where noted otherwise. Multivariable models included all factors significantly associated with folic acid-containing supplement use in the univariate model; factors whose global association was not significant were excluded and denoted with n.s. Separate models were run for income and education, and for province/territory of residence and region of residence to avoid collinearity. Bold indicates significant differences (p<0.05) from the reference group after adjustment for multiple comparisons. Superscript E indicates that values should be interpreted with caution due to a wide CV (16.6-33.3%), while estimates replaced by an F are suppressed due to low sample size (<10) and/or a CV ≥33.3%.

2 Sample sizes refer to those who answered either yes or no. Proportions may not add to 100%/the full sample size because some responses were do not know or refuse to answer, or because question was not included for all cycles/and/or provinces/territories as specified below.

3 Total household income before taxes adjusted for household size (income/square root of size)(2). Income available in this format only for 2017/18 cycle.

4 Categorized as Atlantic [New Brunswick, Newfoundland and Labrador, Nova Scotia, Prince Edward Island], Central [Ontario and Quebec], Northern [Yukon, Northwest Territories, Nunavut], Western [Alberta, British Columbia, Manitoba, Saskatchewan].

5 Module was optional content for 2015-16 for NFLD and Labrador, Ontario, module not included for Yukon.

6 Module was core content for 2015/16 only.

7 Question included as optional content in the 2015/16 survey only.
